# Plasma neurofilament light links mortality to neuroaxonal injury beyond clinical decline and Alzheimer disease plasma biomarkers: a multi-cohort study of oldest old individuals

**DOI:** 10.64898/2026.10.17.26365798

**Authors:** Linda M.C. Lorenz, Sietske A.M. Sikkes, Martijn W. Heymans, Marc Hulsman, Niccolò Tesi, Mariam Gouda, Marieke J.I. Graat, Myke E. van der Hoorn, Dominique A.H. Daatselaar, Kaare Christensen, Mikael Thinggaard, Charlotte E. Teunissen, Henne Holstege

## Abstract

Blood-based neurofilament light (NfL) is associated with all-cause mortality, but whether this reflects neuroaxonal injury or overlaps with cognitive and functional decline remains unclear. We followed 559 individuals aged 92–107 from three oldest-old cohorts for up to 16 years: cognitively healthy Dutch centenarians and two population-based Danish cohorts. Plasma biomarkers (Aβ_42/40_, Aβ_40_, Aβ_42_, pTau-181, pTau-181/Aβ_42_, NfL, and GFAP; Dutch cohort; NfL, Danish cohorts), cognition, and activities of daily living (ADL) were evaluated against mortality. NfL was the only biomarker consistently associated with mortality (HR 1.21–1.38), whereas AD-specific biomarkers did not predict mortality despite reflecting *post-mortem* AD neuropathology. NfL remained associated with mortality independently of cognition and ADL; ADL mediated 13–21% across cohorts and cognition 28%, only where cognitive impairment was widespread. Mortality at extreme age is more closely linked to neuroaxonal injury than to upstream AD pathology, with NfL capturing biological vulnerability beyond cognitive and functional decline.

---

Life expectancy at age 80 has risen steadily in high-income countries, resulting in a growing number of individuals reaching ages at which mortality is increasingly shaped by age-related disease.^1,2^ Alzheimer Disease (AD), the leading cause of dementia^3,4^ is associated with increased mortality.^5–7^ AD is characterized by extracellular amyloid-β (Aβ) plaques, intracellular neurofibrillary tau tangles (NFT), neuroinflammation and neurodegeneration.^8^ Blood-based biomarkers now allow these neuropathological processes to be tracked *in vivo*, including AD-specific markers such as the Aβ_42/40_ ratio and phosphorylated tau-181 (pTau-181), as well as broader markers of neurodegeneration neurofilament light chain (NfL) and glial fibrillary acidic protein (GFAP).^9^ Among the oldest old, however, survival varies markedly despite similar chronological ages, suggesting substantial biological heterogeneity. Blood-based biomarkers therefore provide an opportunity to determine whether mortality at extreme age is more closely linked to Alzheimer pathology, neuroaxonal injury, or the clinical consequences of brain aging.

We previously demonstrated that, in cognitively healthy centenarians from the Dutch 100-plus Study, plasma Aβ_42_ and particularly pTau-181 closely reflect post-mortem AD neuropathology.^10^ This raises a fundamental question: Does mortality at extreme age primarily reflect AD pathology itself, or the downstream neuroaxonal injury and clinical decline that may result from it? If AD pathology contributes directly to mortality, we hypothesize that AD-specific plasma biomarkers should predict survival. Conversely, if mortality is more closely linked to neuroaxonal injury, biomarkers such as NfL would be expected to better discriminate survival.

Plasma NfL,^11,12^ cognitive performance (Mini-Mental State Examination; MMSE) and activities of daily living (ADL) are well-established predictors of mortality across adulthood and into extreme old age,^13–16^ while GFAP has also been associated with mortality in some populations.^17^ However, to what extent NfL simply reflects cognitive and functional impairment, or captures additional biological vulnerability is currently unclear. Also, it remains unknown whether AD-specific plasma biomarkers provide prognostic information beyond these established clinical and neurodegeneration markers. A recent study in cognitively impaired Japanese centenarians, reported that NfL, but neither the Aβ_42/40_ ratio nor pTau-181, predicted mortality, consistent with the possibility that neuroaxonal injury is more closely linked to survival than upstream AD pathology.^12^ However, that study did not test whether the NfL-mortality association was independent of, or mediated through, cognition and ADL functioning, nor whether this relationship generalizes across oldest-old cohorts with different cognitive and functional impairment.

To address these questions, we studied three complementary cohorts of the oldest old. The cognitively healthy Dutch 100-plus Study, in which the full panel of AD-related plasma biomarkers was available, allowed us to distinguish whether mortality is more closely linked to Alzheimer’s pathology or neuroaxonal injury. Two population-based Danish cohorts, a centenarian cohort (1915 West Birth Cohort) and a nonagenarian cohort (1905 Birth Cohort) previously described by Kaeser et al. (2021)^11^ then enabled us to determine whether the NfL-mortality association generalized across different levels of cognitive and functional impairment and whether it remained independent of, or was mediated through, cognition and ADL functioning.

## RESULTS

### Characteristics of three complementary oldest-old cohorts

As ADL functioning was assessed using different instruments (Barthel Index, Dutch; Katz Index, Danish), scores were categorized into cohort-specific tertiles (lower, middle, higher) for descriptive purposes only and are not directly comparable across cohorts (**Extended Data Table 1**).

#### Dutch 100-plus Study (self-reported cognitively healthy centenarians)

The cognitively healthy Dutch centenarian cohort (N=255) had a median baseline age of 101.3 years (interquartile range [IQR]: 100.3 to 102.4) and was followed for a median of 1.42 years, during which most participants died (233 of 255, 91%) and 22 (9%) were censored (453.6 person-years; mortality rate 51.4 per 100 person-years). Baseline cognitive and ADL functioning were relatively high for this age (median MMSE 26/30; median BI 15/20). Decedents and censored participants did not differ in sex, cognitive performance, or ADL functioning (all *P≥*0.470), though decedents were slightly older at blood collection (*P*=0.038). Males had slightly better cognitive performance and higher ADL functioning than females (all *P*S 0.014; **Table 1**).

**Table 1.**
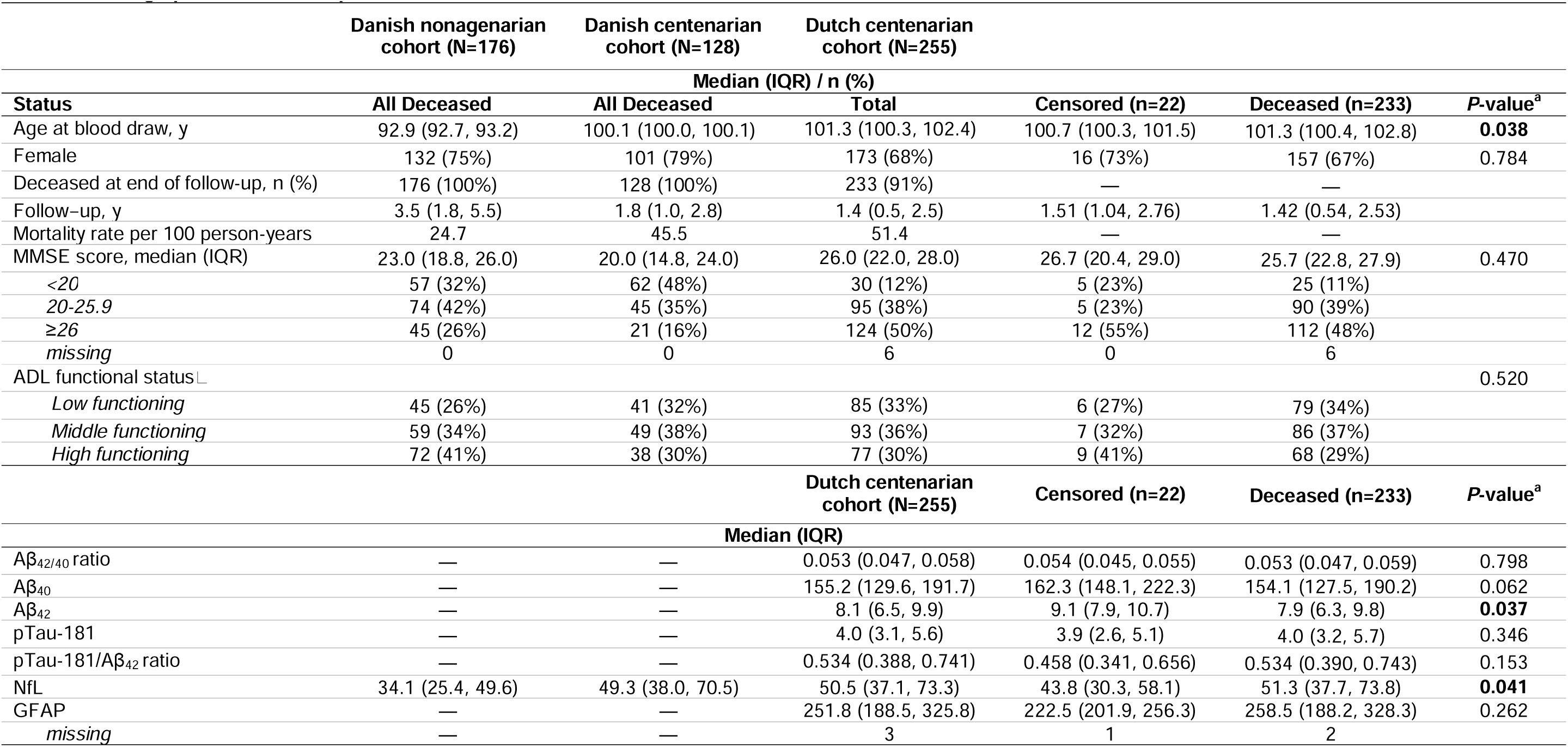
Demographic, clinical, and plasma biomarker characteristics.

#### Danish 1915 West Birth Cohort (population-based centenarians)

The population-based Danish centenarian cohort (N=128) had a median baseline age of 100.1 years (IQR: 100.0 to 100.1) and median survival of 1.81 years (281.5 person-years). All were deceased by end of follow-up (mortality rate 45.5 per 100 person-years). Consistent with population-based inclusion, cognitive performance was lower than in the selected Dutch cohort (median MMSE 20/30). Danish biomarker analyses were restricted to NfL (median 49.3 pg/mL). Males had higher ADL functioning than females (*P*=0.040; **Table 1**).

#### Danish 1905 Birth Cohort (population-based nonagenarians)

The population-based Danish nonagenarian cohort (N=176) had a median baseline age of 92.9 years (IQR 92.7–93.2) and median survival of 3.51 years (IQR 1.8–5.5), contributing 713.9 person-years (mortality rate 24.7 per 100 person-years). Cognitive performance was intermediate between the centenarian cohorts (median MMSE 23.0, IQR 18.8–26.0), and NfL was lower than in either centenarian cohort (median 34.1 pg/mL, IQR 25.4–49.6). Full characteristics for all three cohorts appear in **Table 1**.

### Distinct predictors of mortality across cohorts: robustness of plasma NfL, with ADL and MMSE varying by cohort

Unadjusted survival trajectories are shown in **Figure 1** and **Extended Data Figure 1**. To evaluate the independent prognostic value of each biomarker and clinical measure, we fitted stepwise Cox proportional-hazards models successively adjusting for age and sex in Model 1; age, sex, and MMSE in Model 2; age, sex, and ADL functioning in Model 3; age, sex, MMSE, and ADL functioning in Model 4; and age, sex, MMSE, ADL functioning, and NfL in Model 5, and where appropriate, we added the remaining AD-specific biomarkers in Model 6 (**Table 2**). **Figure 2** and **Extended Data Figure 2** show a single fully adjusted model per predictor (the final, mutually adjusted estimate). Multivariable fractional polynomial analyses of all biomarkers and clinical predictors are shown in **Extended Data Table 3**.

**Figure 1.**
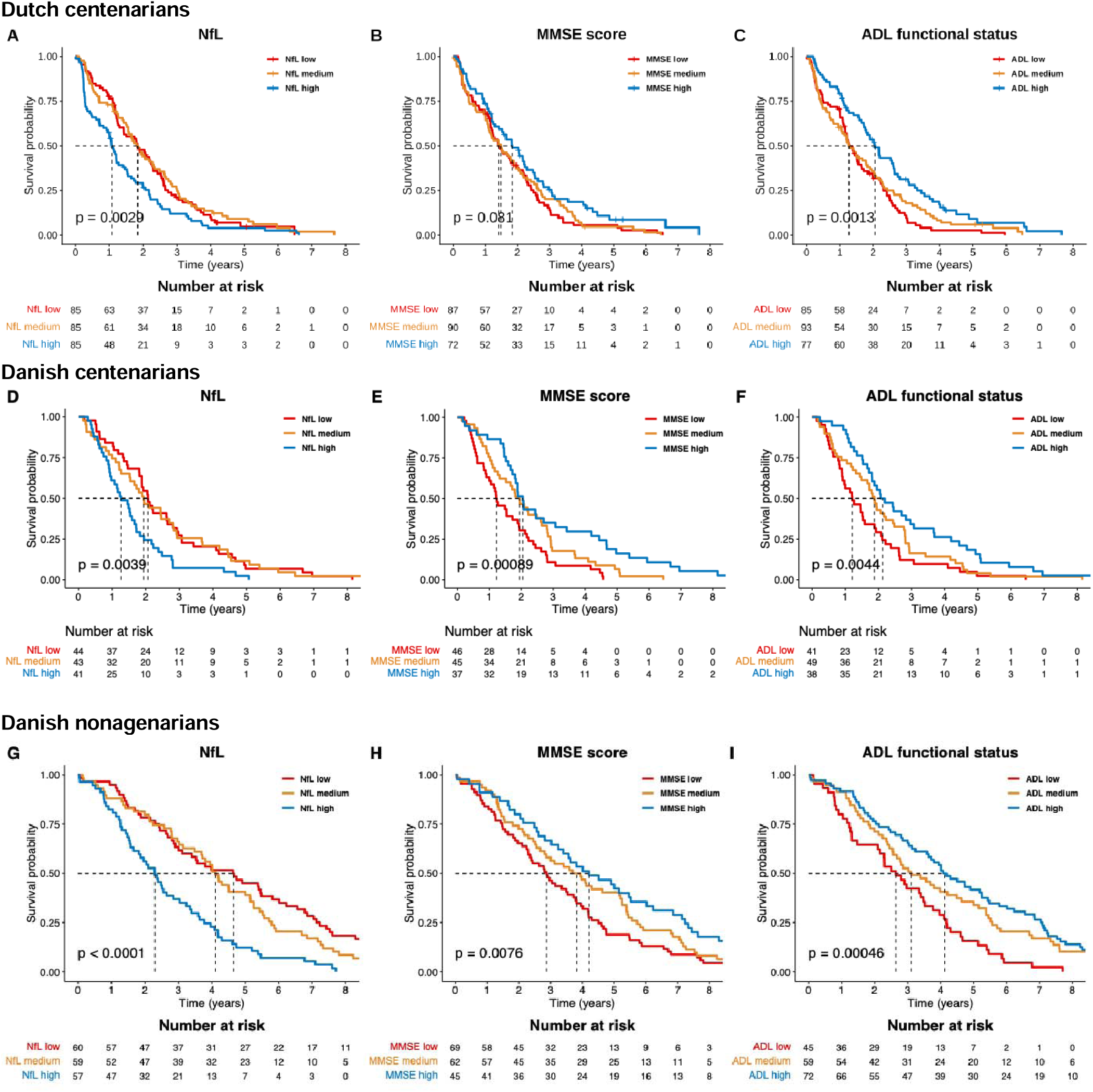
Kaplan-Meier survival curves for plasma NfL, MMSE score, and ADL functioning. *Abbreviations*: ADL, activities of daily living; BI, Barthel Index; Katz, Katz Index; MMSE, Mini-Mental State Examination; NfL, neurofilament light chain. Survival probability (y-axis) from blood collection to death (x-axis, years) is shown for participants stratified into cohort-specific tertiles (low, medium, high). ADL was assessed with the Barthel Index (Dutch) and the Katz Index (Danish). *P*-values are from global log-rank tests across the three tertiles.

**Figure 2.**
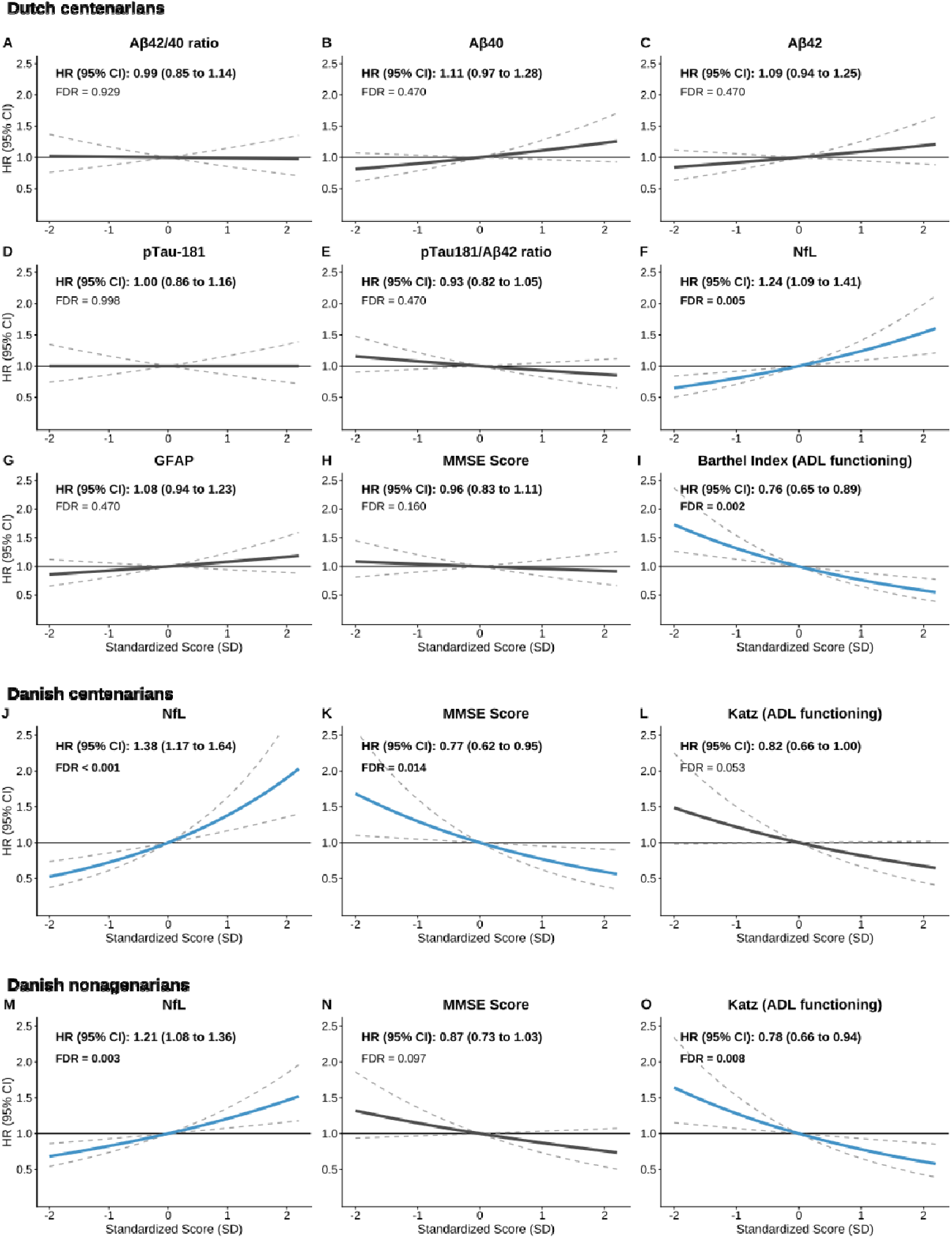
Adjusted hazard ratio curves for all-cause mortality across plasma biomarkers, cognitive, and ADL functioning. *Abbreviations*: ADL, activities of daily living; Aβ, amyloid-β; BI, Barthel Index; CI, confidence interval; GFAP, glial fibrillary acidic protein; HR, hazard ratio; Katz, Katz Index; MMSE, Mini-Mental State Examination; NfL, neurofilament light chain; pTau-181, phosphorylated tau-181; SD, standard deviation. Solid lines show the HR across the standardized predictor score (x-axis); dashed lines are 95%CIs. Blue indicates statistical significance (*FDR*<0.05 for plasma biomarkers; *P*<0.05 for the NfL-adjusted MMSE and ADL); grey indicates non-significant. Plasma biomarkers were adjusted for age, sex, MMSE, and ADL (BI in the Dutch cohort, Katz in the Danish cohorts); MMSE was adjusted for age, sex, ADL, and NfL; ADL was adjusted for age, sex, MMSE, and NfL.

**Table 2.** Cox proportional hazards models for all-cause mortality.

| Predictor | Models 1<br>(individual) |  | Models 2<br>(individual) |  | Models 3<br>(individual) |  | Models 4<br>(individual) |  | Models 5<br>(individual) |  | Model 6<br>(simultaneous) |  |
| --- | --- | --- | --- | --- | --- | --- | --- | --- | --- | --- | --- | --- |
| | Adj. age, sex | | Adj. age, sex,<br>MMSE | | Adj. age, sex,<br>ADL functioning | | Adj. age, sex,<br>MMSE,<br>ADL functioning | | Adj. age, sex,<br>MMSE,<br>ADL functioning, NfL | | Adj. age, sex, MMSE,<br>ADL functioning,<br>all biomarkers<br>(Aβ<sub>42/40</sub> ratio, Aβ<sub>40</sub>, Aβ<sub>42</sub>,<br>pTau-181, pTau-181/Aβ<sub>42</sub>,<br>NfL, and GFAP) | |
|  | HR (95%CI) | FDR | HR (95%CI) | FDR | HR (95%CI) | FDR | HR (95%CI) | FDR | HR (95%CI) | FDR | HR (95%CI) | P |
| **Dutch centenarian cohort** |  |  |  |  |  |  |  |  |  |  |  |  |
| *n*=255 |  |  |  |  |  |  |  |  |  |  |  |  |
| Aβ<sub>42/40</sub> ratio | 1.00 (0.87, 1.15) | 0.986 | 1.01 (0.88, 1.16) | 0.950 | 0.98 (0.85, 1.12) | 0.903 | 0.99 (0.85,1.14) | 0.929 | 0.98 (0.85, 1.13) | 0.909 | 1.19 (0.75, 1.89) | 0.458 |
| Aβ<sub>40</sub> | 1.09 (0.95, 1.25) | 0.468 | 1.10 (0.96, 1.27) | 0.468 | 1.10 (0.96, 1.27) | 0.468 | 1.11 (0.97,1.28) | 0.470 | 1.07 (0.93, 1.24) | 0.537 | 1.56 (0.75, 3.25) | 0.236 |
| Aβ<sub>42</sub> | 1.08 (0.94, 1.24) | 0.468 | 1.10 (0.96, 1.27) | 0.468 | 1.07 (0.93, 1.23) | 0.537 | 1.09 (0.94,1.25) | 0.470 | 1.04 (0.90, 1.20) | 0.784 | 0.63 (0.31, 1.27) | 0.199 |
| pTau-181 | 1.01 (0.87, 1.18) | 0.950 | 1.02 (0.88, 1.18) | 0.949 | 1.00 (0.87, 1.16) | 0.986 | 1.00 (0.86,1.16) | 0.998 | 0.94 (0.81, 1.10) | 0.662 | 1.03 (0.74, 1.42) | 0.882 |
| pTau-181/Aβ<sub>42</sub> ratio | 0.93 (0.82, 1.05) | 0.468 | 0.92 (0.81, 1.05) | 0.468 | 0.94 (0.83, 1.06) | 0.537 | 0.93 (0.82,1.05) | 0.470 | 0.92 (0.81, 1.05) | 0.468 | 0.86 (0.62, 1.21) | 0.391 |
| NfL | 1.29 (1.15, 1.45) | **<0.001** | 1.26 (1.12, 1.43) | **0.002** | 1.25 (1.11, 1.41) | **0.003** | 1.24 (1.09,1.41) | **0.005** | — | — | 1.24 (1.08, 1.42) | **0.003** |
| GFAP | 1.11 (0.97, 1.27) | 0.468 | 1.08 (0.94, 1.24) | 0.468 | 1.08 (0.94, 1.23) | 0.468 | 1.08 (0.94,1.23) | 0.470 | 1.03 (0.89, 1.19) | 0.902 | 1.05 (0.90, 1.23) | 0.537 |
| MMSE | 0.84 (0.73, 0.96) | **0.043** | — | — | 0.95 (0.81, 1.10) | 0.663 | — | — | — | — | 0.99 (0.85, 1.15) | 0.887 |
| ADL functioning | 0.73 (0.63, 0.83) | **<0.001** | 0.74 (0.64, 0.87) | **0.002** | — | — | — | — | — | — | 0.75 (0.64, 0.88) | **<0.001** |
| **Danish centenarian cohort** |  |  |  |  |  |  |  |  |  |  |  |  |
| *n*=128 |  |  |  |  |  |  |  |  |  |  |  |  |
| NfL | 1.50 (1.27, 1.77) | **<0.001** | 1.38 (1.16, 1.64) | **<0.001** | 1.45 (1.23, 1.71) | **<0.001** | 1.38 (1.17, 1.64) | **<0.001** | — | — | — | — |
| MMSE | 0.67 (0.56, 0.81) | **<0.001** | — | — | 0.73 (0.59, 0.89) | **0.003** | — | — | — | — | — | — |
| ADL functioning | 0.73 (0.61, 0.87) | **0.001** | 0.82 (0.67, 1.01) | 0.062 | — | — | — | — | — | — | — | — |
| **Danish nonagenarian cohort** |  |  |  |  |  |  |  |  |  |  |  |  |
| *n*=176 |  |  |  |  |  |  |  |  |  |  |  |  |
| NfL | 1.22 (1.10, 1.36) | **<0.001** | 1.22 (1.09, 1.37) | **0.001** | 1.21 (1.08, 1.36) | **0.003** | 1.21 (1.08, 1.36) | **0.003** | — | — | — | — |
| MMSE | 0.79 (0.68, 0.92) | **0.003** | — | — | 0.86 (0.73, 1.02) | 0.084 | — | — | — | — | — | — |
| ADL functioning | 0.71 (0.61, 0.84) | **<0.001** | 0.76 (0.64, 0.91) | **0.005** | — | — | — | — | — | — | — | — |

#### Dutch centenarian cohort (self-reported cognitively healthy)

NfL was the only biomarker consistently associated with mortality (**Figure 2**; **Extended Data Figure 2**). In the age- and sex-adjusted model, higher NfL predicted increased hazard (Model 1; HR 1.29, 95%CI 1.15–1.45; *FDR<*0.001). This NfL log-hazard was reduced by 9% when we adjusted additionally for MMSE (Model 2), while it was reduced by 14% when adjusted by ADL alone (Model 3), and when adjusted by both MMSE and ADL, the NfL association was reduced by 17% (Model 4; HR 1.24, 95%CI 1.09–1.41; *FDR*=0.005; **Extended Data Table 4**). The association between NfL and mortality did not change upon further adjustment for the complete AD-related biomarker panel (Model 6: HR 1.24, 95%CI 1.08–1.42; *P*=0.003). Furthermore, the NfL-mortality association was robust to exclusion of four participants with extreme NfL concentrations (S300 pg/mL; HR 1.30, 95% CI 1.12-1.51; *P*<0.001), indicating that the observed association was not driven by outlying observations. No other biomarker (Aβ_42/40_ ratio, Aβ_40_, Aβ_42_, pTau-181, pTau-181/Aβ_42_ ratio, GFAP) was individually associated with mortality in any model (all *FDR≥*0.468; **Table 2**). With 233 deaths, the cohort had 80% power to detect HR ∼1.21 per 1-SD (**Extended Data Table 2**), indicating that any mortality associations of the AD-specific biomarkers are non-existent or at least substantially smaller than those observed for NfL. Among the clinical measures, ADL functioning remained independently associated with mortality after adjustment for NfL and MMSE (HR 0.76, 95% CI 0.65 to 0.89; *P*=0.002), whereas the association between MMSE and mortality was mostly attenuated by both ADL and NfL and was no longer significant after mutual adjustment (HR 0.96, 95% CI 0.83 to 1.11; *P*=0.160).

#### Danish 1915 West Birth Cohort (population-based centenarians)

The association between NfL and mortality was also evident in the population-based Danish centenarians (**Figure 2**; **Extended Data Figure 2**). Higher NfL predicted increased hazard (Model 1 HR 1.50, 95%CI 1.27–1.77; *FDR*<0.001). This association remained stable when we adjusted for MMSE and ADL functioning: adding MMSE (Model 2) alone reduced the NfL log-hazard by 20%, ADL alone by 8% (Model 3), and both together by 20% (Model 4 HR 1.38, 95%CI 1.17–1.64; *FDR*<0.001). The association was robust to exclusion of the single participant with an extreme NfL concentration (≥300 pg/mL; HR 1.31, 95%CI 1.10-1.56; *P* =0.003). With 128 deaths, the cohort had 80% power to detect HR ∼1.28 per 1-SD (**Extended Data Table 2**), and the observed HR of 1.50 lay well above this threshold.

Unlike the cognitively healthy Dutch centenarians, cognitive performance emerged as the strongest clinical predictor of mortality in the mostly declined Danish centenarians. MMSE remained independently associated with mortality after adjustment for NfL and ADL functioning (HR 0.77, 95%CI 0.62 to 0.95; *P*=0.014), whereas the association between ADL and mortality was largely attenuated by MMSE and was no longer significant after mutual adjustment (HR 0.82, 95%CI 0.66 to 1.00; *P*=0.053).

#### Danish 1905 Birth Cohort (population-based nonagenarians)

The association between NfL and mortality also replicated in the population-based Danish nonagenarians, the youngest of the three cohorts and the one with the lowest absolute NfL concentrations (Model 1 HR 1.22, 95% CI 1.10–1.36; *FDR*<0.001; **Table 2**; **Extended Data Figure 2**). Higher NfL concentrations remained associated with increased mortality after adjustment for MMSE and ADL functioning (Models 2 and 3), and for the fully adjusted Model 4 (HR 1.21, 95% CI 1.08–1.36; *FDR*=0.003). The association was strengthened after excluding the single participant with NfL S300 pg/mL (HR 1.39, 95% CI 1.19-1.62; *P*<0.001). With 176 deaths, the study had 80% power to detect hazard ratios of approximately 1.24 per SD; the observed HR of 1.22 sat close to this threshold but remained significant.

As in the cognitively healthy Dutch centenarians, ADL functioning emerged as the strongest clinical predictor of mortality in these more declined nonagenarians. ADL functioning remained independently associated with mortality after adjustment for NfL and MMSE (HR 0.78, 95% CI 0.66–0.94; *P*=0.008), whereas the association between MMSE and mortality was largely attenuated by ADL and was no longer statistically significant after mutual adjustment (HR 0.87, 95% CI 0.73–1.03; *P*=0.097). Adjustment for MMSE and ADL reduced the NfL association by only 5%, indicating that the NfL signal remained essentially independent of measured cognitive and ADL impairment.

### Most of the NfL-mortality association is independent of cognition and ADL functioning

Because NfL, MMSE, and ADL functioning were each associated with mortality, we next asked whether the prognostic value of NfL primarily reflected cognitive or ADL decline. Mediation analyses (adjusted for age and sex) showed that cognition and ADL mediated only a minority of the NfL association with mortality (**Figure 3**; **Extended Data Table 5**).

**Figure 3.**
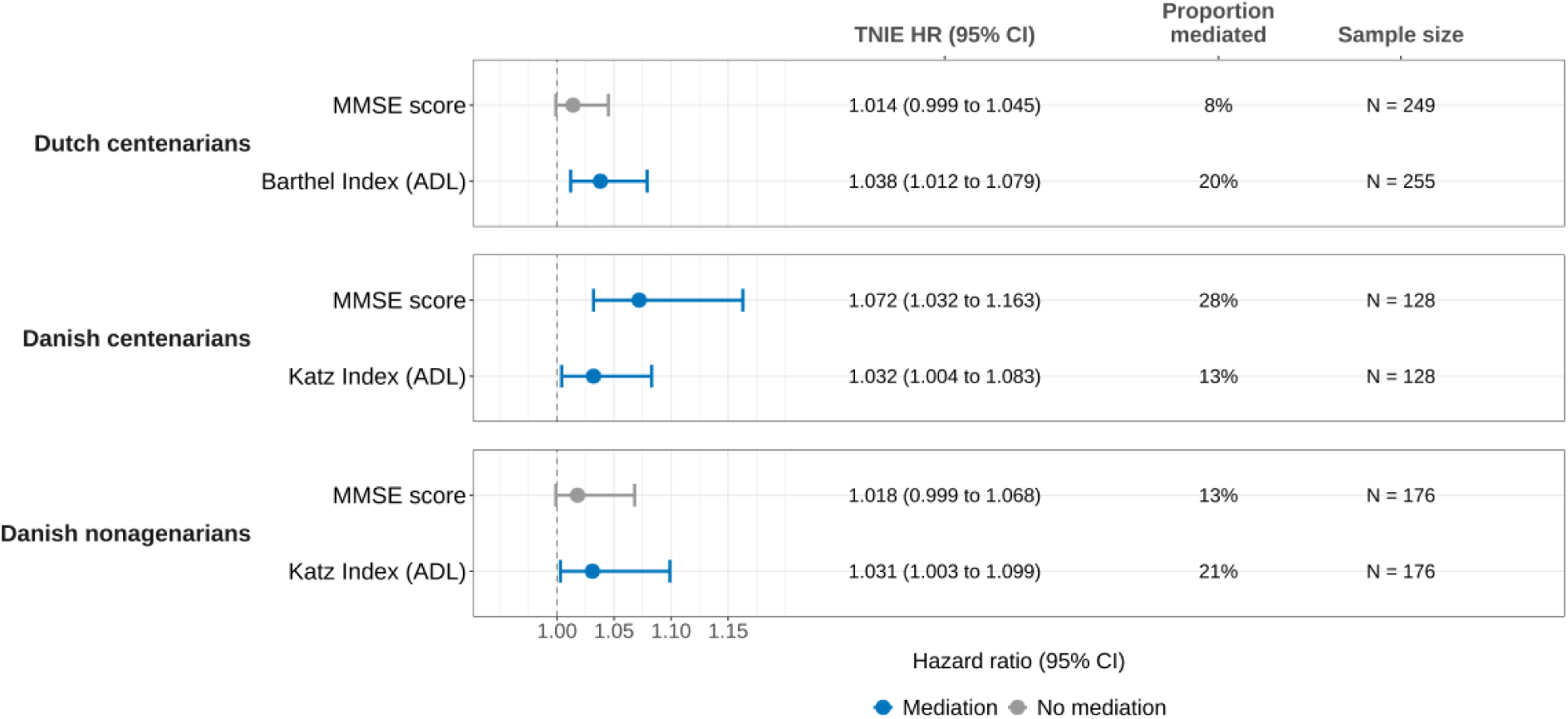
Mediated (indirect) effect of plasma NfL on all-cause mortality through cognitive performance (MMSE) and ADL functioning (Barthel/Katz Index), in (A) Dutch centenarians, (B) Danish centenarians, and (C) Danish nonagenarians. *Abbreviations*: ADL, activities of daily living; CI, confidence interval; HR, hazard ratio; MMSE, Mini-Mental State Examination; NfL, neurofilament light chain; TNIE, total natural indirect effect. HRs (95% CI) are total natural indirect effects (TNIE) for a P75 vs P25 NfL contrast, adjusted for age and sex. Blue indicates evidence of mediation (TNIE 95% CI excludes 1); grey, no evidence.

How much each pathway contributed depended on the cognitive state of the cohort. In the cognitively healthy Dutch centenarians, only ADL functioning significantly mediated the NfL-mortality association, accounting for ∼20% of the total effect. In the cognitively impaired Danish centenarians, both MMSE and ADL pathways were significant, with MMSE accounting for the larger proportion (∼28%) and ADL ∼13%. In the younger Danish nonagenarians, ADL again mediated a significant proportion (∼21%), whereas MMSE did not. ADL functioning was therefore the consistent mediating pathway between NfL and mortality across cohorts, whereas MMSE contributed only where cognitive impairment was widespread.

In all three cohorts, the direct effect of NfL remained the dominant component, indicating that NfL captures biological vulnerability beyond what is reflected by cognition or ADL impairment. No significant mediation was observed for any of the other plasma biomarkers (**Extended Data Table 6**).

### NfL adds prognostic value beyond cognitive and ADL functioning scales

Nested Cox models tested whether NfL improved mortality prediction beyond clinical measures of cognitive and ADL status. In all three cohorts, adding NfL improved model fit beyond MMSE and ADL functioning alone (all *P*<0.001; **Extended Data Table 7**). In the Dutch centenarians, where the full biomarker panel was available, NfL was the only plasma biomarker that improved model performance (ΔAIC=-10.6; all others S-0.2) and the best model combined NfL with ADL functioning (ΔAIC=-23.3), MMSE adding nothing further (ΔAIC=-21.5). The clinical predictor contributing most to mortality prediction differed by cohort: ADL functioning contributed most in the cognitively healthy Dutch centenarians and Danish nonagenarians, whereas MMSE contributed more in the cognitively impaired Danish centenarians. Across all three cohorts, NfL consistently improved prediction, supporting its non-redundant prognostic value beyond measured cognitive and ADL status.

### The prognostic value of NfL increases over longer survival horizons

To quantify the clinical impact of NfL, we estimated restricted mean survival time (RMST), the mean number of years expected to survive within a fixed window. RMST was estimated with age, sex, and ADL functioning held constant (**Figure 4A**; **Extended Data Table 8**). Across all three cohorts, the survival difference associated with higher NfL increased with longer follow-up. Thus, rather than imposing a fixed reduction in life expectancy, the prognostic effect of NfL accumulated over time.

**Figure 4A.**
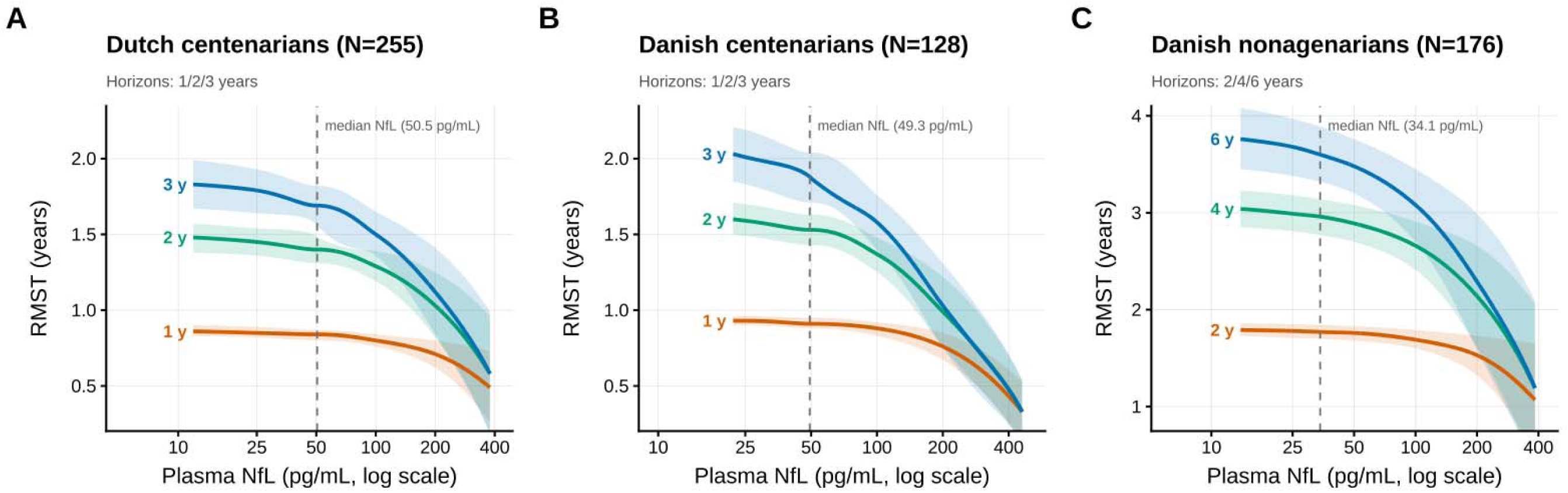
Restricted mean survival time (RMST) as a function of plasma NfL. *Abbreviations*: ADL, activities of daily living; BI, Barthel Index; CI, confidence interval; Katz, Katz Index; MMSE, Mini-Mental State Examination; NfL, neurofilament light chain; RMST, restricted mean survival time; SD, standard deviation. (Plot A) Dutch centenarians (N = 255), (Plot B) Danish centenarians (N = 128) and (Plot C) Danish nonagenarians (N = 176). In each panel, solid lines show predicted RMST (y-axis, years) across plasma NfL (x-axis, pg/mL, log scale), one line per survival-time horizon (1, 2 and 3 years for the centenarian cohorts; 2, 4 and 6 years for the nonagenarian cohort), and shaded areas are 95% CIs. The dashed vertical line marks each cohort’s median plasma NfL. RMST was predicted from flexible parametric survival models with age, sex and ADL functioning held at cohort means.

For example, in the Dutch centenarian cohort an individual at the median NfL (50.5 pg/mL) had an estimated 70%, 42%, and 19% probability of surviving at least 1, 2, and 3 years, respectively. An individual with roughly double that level (100 pg/mL) had 63%, 33%, and 13% probabilities over 1-, 2-, and 3-year horizons. Expressed as restricted mean survival time, this shortened expected survival by roughly 0.4, 1.3, and 2.3 months over the same horizons. Furthermore, the penalty did not depend on ADL functioning: the NfL effect was comparable at higher and lower ADL. Similar patterns were observed in both Danish cohorts, where higher NfL likewise increasingly separated survival trajectories over their respective projection horizons (**Figure 4A**; **Extended Data Table 8**).

Across all cohorts, the effect of NfL was largely independent of ADL functioning, with all NfL×ADL interactions non-significant (**Extended Data Table 10**), and was evident irrespective of baseline ADL functioning. Per-standard-deviation RMST effects were smallest in the cognitively healthy Dutch centenarians and larger in the two Danish cohorts (**Figure 4B**), consistent with stronger mortality associations in cohorts with greater cognitive and ADL impairment. Together, these findings indicate that NfL captures biological vulnerability that influences survival progressively over time, rather than merely identifying individuals at imminent risk of death.

**Figure 4B.**
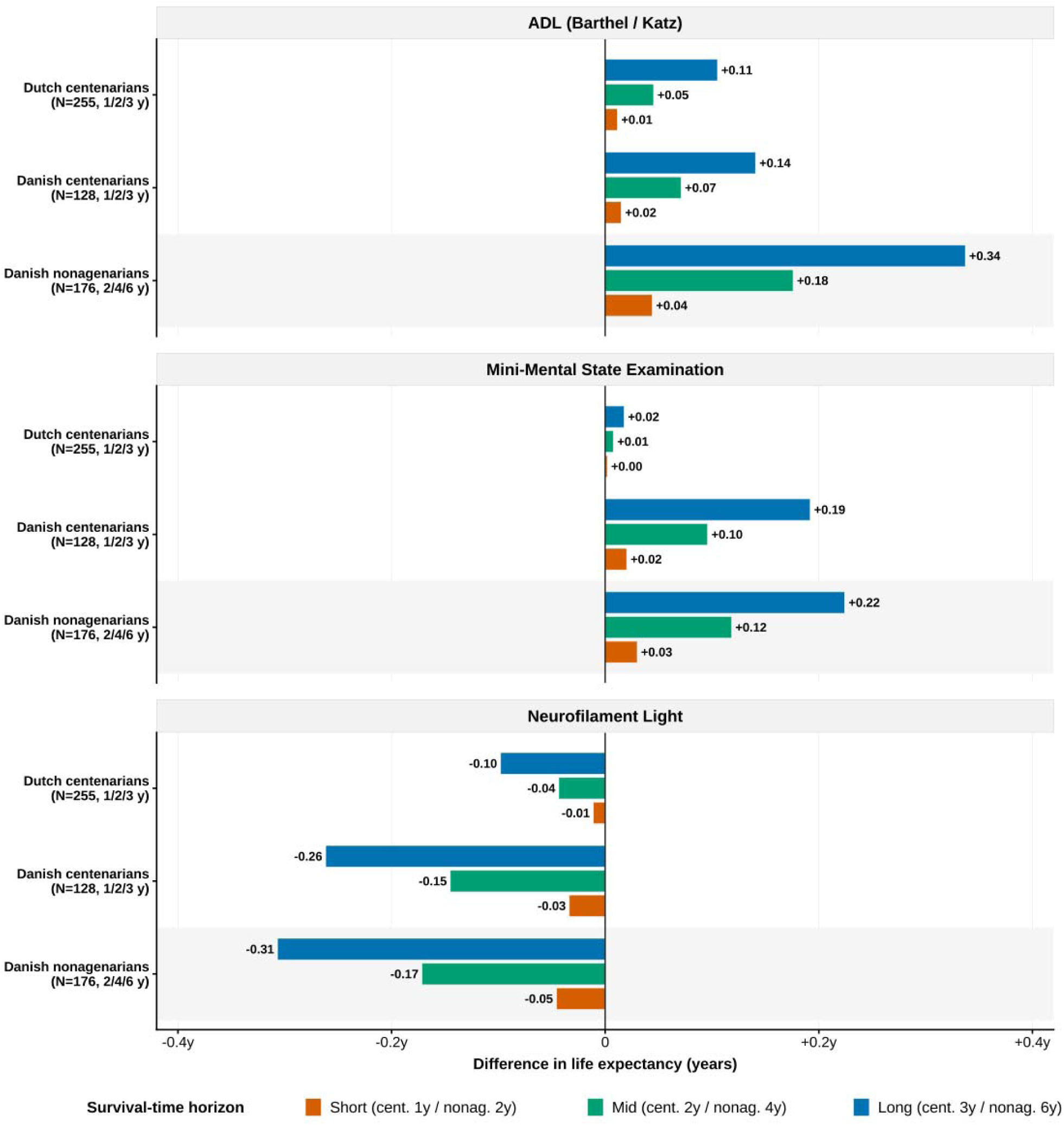
The impact of NfL, MMSE, and ADL on remaining life expectancy across three oldest-old cohorts. *Abbreviations*: ADL, activities of daily living; BI, Barthel Index; CI, confidence interval; Katz, Katz Index; MMSE, Mini-Mental State Examination; NfL, neurofilament light chain; SD, standard deviation. Bars show the difference in life expectancy (years) per 1-SD increase in ADL, MMSE, and NfL; higher ADL or MMSE lengthens and higher NfL shortens expected survival. Horizons were 1, 2, and 3 years (centenarian cohorts) and 2, 4, and 6 years (Danish nonagenarians); ADL was the BI (Dutch) and the Katz Index (Danish).

## DISCUSSION

We investigated whether mortality at extreme age is more closely associated with AD-specific pathology, general neurodegeneration, or clinical measures of cognitive and ADL decline by studying plasma biomarkers, cognition, and ADL functioning in cognitively healthy centenarians from the Dutch 100-plus Study, together with two population-based Danish cohorts of nonagenarians and centenarians. We found that first, NfL was the only plasma biomarker consistently associated with mortality, whereas AD-specific plasma biomarkers were not. Second, this association remained largely independent of cognition and ADL functioning, with only a minority mediated through these clinical measures. Third, NfL improved mortality prediction beyond cognition and ADL particularly over longer follow-up periods.

Plasma NfL predicts all-cause mortality from middle age onward and into extreme age.^11,17–25^ We extend this evidence by showing that the NfL-mortality association is not merely a proxy for observable cognitive or ADL impairment: across all cohorts only ∼13–21% was mediated through ADL functioning, and ∼28% by cognitive performance only in the most cognitively impaired cohort. By comparison, in the two cognitively healthier Dutch centenarian- and Danish nonagenarian cohorts, cognitive performance mediated only 8% and 13% of the association, and neither reached statistical significance. Nevertheless, NfL remained associated with mortality independently from MMSE and ADL, indicating that plasma NfL captures biological vulnerability that is not reflected by conventional measures of cognition and ADL status^16,26,27^.

NfL was not only independently associated with mortality but was also the only biomarker that consistently improved mortality prediction beyond cognition and ADL functioning. In the Dutch cohort, where the full biomarker panel was available, no AD-specific biomarker improved model performance after accounting for NfL. Moreover, the survival difference associated with NfL accumulated over longer follow-up. Together, these findings indicate that NfL captures biological vulnerability that influences survival progressively over time, rather than merely identifying individuals at imminent risk of death.

The absence of independent associations for Aβ_42/40_ ratio, Aβ_42_, pTau-181 or GFAP^17^ is biologically informative.^12,24^ Although Aβ_42_, and especially pTau-181, closely reflect *post-mortem* Aβ and tau neuropathology in the same Dutch centenarians,^10^ they did not predict mortality. Our findings in the oldest-old closely align with a study reporting largely null associations for AD-specific plasma biomarkers^12^ which shows that neurodegeneration markers outperform amyloid and tau biomarkers in predicting mortality. Together, this suggests that mortality across adulthood and into extreme age is more closely aligned with neuroaxonal injury than with AD pathology. Our results are therefore in line with the view that NfL should be viewed less as an Alzheimer disease biomarker than as an integrative marker of cumulative neuroaxonal injury arising from multiple age-related processes and the broader biological vulnerability that ultimately limits survival.

The biological meaning of a given NfL concentration is likely to depend on an individual’s resilience and stage of biological aging. Individuals who survive to exceptional ages represent a survivorship-selected population in whom moderate neuroaxonal injury may be better tolerated than in younger or less resilient individuals.^21,22^ Accordingly, the same NfL concentration may reflect different degrees of biological vulnerability depending on the remaining cognitive and ADL reserve, potentially explaining why NfL predicts mortality throughout adulthood while its overlap with cognition and ADL varies across cohorts.

This study has several strengths, including prospective follow-up of up to 16 years across three cohorts spanning different levels of cognitive and ADL impairment together with same-day blood sampling and clinical assessment, minimizing confounding by interim events. Several limitations should be considered. First, pre-analytical conditions were not fully standardized because blood was collected in homes and care-facilities and residual confounding by factors influencing biomarker concentrations such as renal insufficiency, inflammation and subclinical vascular disease cannot be excluded.^28–31^ Second, while the AD-specific biomarker analyses were restricted to the Dutch cohort, this dataset confirmed the absence of any association of plasma AD-specific biomarker concentrations with mortality as was previously shown in other studies.^12,17,24^

In conclusion, plasma NfL robustly predicts all-cause mortality across oldest old cohorts and provides prognostic information beyond cognition and ADL functioning. The prognostic value of NfL was most evident over extended follow-up, where higher NfL increasingly separated survival trajectories. In contrast, AD-specific Aβ and tau biomarkers did not predict mortality, despite reflecting *post-mortem* AD-neuropathology. Together, these findings identify neuroaxonal injury, rather than upstream Alzheimer’s pathology, as the biological process most closely linked to mortality at extreme age and position plasma NfL as an integrative marker of biological vulnerability.

## ONLINE METHODS

### Cohort characteristics

Detailed descriptions of recruitment, collection procedures, and cohort characteristics have been published previously^11,32^ and are available in **Extended Methods**.

#### Dutch centenarian cohort (100-plus Study)

The 100-plus Study is an ongoing, prospective cohort that recruits centenarians who self-report as cognitively healthy at inclusion, a status confirmed by proxy.

#### Danish cohorts (1915 West birth cohort; 1905 birth cohort)

The Danish cohorts are two population-based birth cohort studies, the 1915 West birth cohort (centenarians) and the 1905 birth cohort (nonagenarians); full inclusion criteria are detailed in the **Extended Methods**.

### Blood collection and Simoa plasma biomarker measurements

#### Dutch centenarian cohort

Cross-sectional non-fasting peripheral blood samples were collected from 255 Dutch centenarians (100-plus Study; 173 females, 68%; median age 101.3 years, IQR: 100.3–102.4) using established protocols.^32^ Plasma concentrations of Aβ_40_, Aβ_42_, pTau-181, NfL, and GFAP were measured using Single Molecule Array (Simoa) technology on a Simoa HD-X analyzer (Quanterix, Billerica, MA, USA). All assays were performed by personnel blinded to clinical and demographic information.

#### Danish cohorts

Cross-sectional non-fasting peripheral blood samples were collected from 128 Danish centenarians (Danish 1915 West birth cohort; 101 females, 79%; median age 100.1 years, IQR: 100.0–100.1) and 176 nonagenarians (Danish 1905 birth cohort; 132 females, 75%; median age 92.9 years, IQR: 92.7– 93.2) using established protocols.^10^ In contrast to the full biomarker panel measured in the Dutch cohort, only plasma NfL concentrations were available for the Danish cohorts, determined using Simoa technology (Quanterix) at the Hertie Institute for Clinical Brain Research, University of Tübingen, as previously described by Kaeser et al. (2021).^11^ Sample processing, assay kits, and quality control are detailed in the Extended Methods; raw biomarker distributions are shown in **Extended Data Figure 3**.

### Demographic data and measurement instruments

#### Dutch centenarian cohort

Demographic information (age, sex) of centenarians was collected at baseline. A full description of the demographic information collected is detailed in prior documentation,^32^ and in **Extended Methods.** ADL functioning was assessed using the Dutch version of the Barthel Index (BI) questionnaire.^33^ Global cognitive functioning was assessed using a modified version of the MMSE,^34^ an 11-section screening tool.^35^ Assessments were conducted in participants’ homes or care facilities by trained research personnel. Missing items were imputed using Multiple Imputation by Chained Equations (MICE) package (v3.13.0)^36^ in R. For details regarding imputation see **Extended Methods** and **Extended Data Table 9**.

#### Danish cohorts

Demographic information (age, sex) of the centenarians and nonagenarians was obtained through the Danish Civil Registration System. A full description of the demographic information collected is detailed in prior documentation,^32^ and in **Extended Methods.** ADL functioning was assessed using a modified version of the Katz Index which did not include the evaluation of continence.^37^ Hence, the score only ranged from 0 to 5 and higher scores denote greater ADL functional independence, mirroring the directional orientation of BI used in the Dutch cohort. Global cognitive functioning was assessed using a modified version of the MMSE. Assessments were conducted in participants’ homes or care facilities by trained lay personnel in the Danish 1905 birth cohort and by geriatricians and a nurse in the Danish 1915 West birth cohort.

### Mortality assessment

#### Dutch cohort

Mortality data were collected based on family and caregiver reports until June 2026, at which point the survival times for centenarians not confirmed deceased were administratively censored using their last confirmed date alive. Survival time was defined as the time interval from the date of blood collection to either the date of death or the most recent confirmation of being alive. Of the 255 centenarians included in this study, 7 (3%) were lost to follow-up as of June 2026.

#### Danish cohorts

Mortality data were obtained via linkage with the Danish Civil Registration System using unique identification numbers, ensuring 100% follow-up.

### Ethical oversight

All participants or their legal proxies provided written informed consent, and the studies were conducted in accordance with the Declaration of Helsinki.

#### Dutch centenarian cohort

This study was approved by the Amsterdam UMC Medical Ethics Committee (registration 2016.440).

#### Danish cohorts

The studies of the validation cohorts were approved by the Danish ethical committees (approval nos. S-20140099 and VF20040240), the Danish Data Protection Agency (trial nos. 2016-41-4552 and 2020-522-0256), and the ethics committee of the medical faculty at the University of Tübingen, Germany (project nos. 837/2017BO2 and 838/2017BO2).

### Statistical analysis

Our analyses followed the Strengthening the Reporting of Observational Studies in Epidemiology (STROBE) reporting guidelines. Baseline characteristics were compared between participants who died during follow-up and those who were censored (Dutch cohort), using Mann-Whitney *U* tests for continuous variables and x2 tests for categorical variables. All continuous predictors (plasma biomarkers, MMSE, and ADL scores) were z-standardized within each cohort on their raw scale without prior log-transformation, such that hazard ratios reflect the risk per 1-SD increase relative to the cohort-specific distribution; robustness to extreme biomarker values was confirmed by the NfL outlier exclusion sensitivity analysis described below.

#### Cox proportional-hazards regressions

For the primary survival analysis, Cox proportional-hazards regressions estimated hazard ratios (HRs) and 95% confidence intervals (CIs) per 1-SD increase in z-standardized predictors using the R package ‘survival’^38^ (v.3.8.3). The proportional-hazards assumption was checked with Schoenfeld residuals, and five progressively adjusted models plus biomarker × clinical-function interaction terms were fitted (Extended Methods). Attenuation was quantified as the percentage change in log-hazard coefficients (β) between models, and Kaplan-Meier curves were drawn with the visualization package ‘survminer’^39^ (v.0.5.0). Two sensitivity analyses were performed: excluding participants with plasma NfL S300 pg/mL, and, in the Dutch cohort, additionally adjusting NfL for all other AD-associated plasma biomarkers (Aβ_42/40_ ratio, Aβ_40_, Aβ_42_, pTau-181, pTau-181/Aβ_42_, and GFAP). For significant predictors, multivariable fractional polynomial (MFP) models using the package ‘mfp’^40^ assessed non-linearity, adjusting for age, sex, MMSE, and ADL functioning. A posteriori sensitivity was summarized as the minimum HR per 1-SD increase detectable at 80% power, given the observed deaths per cohort, using an analytical events-based formula (Schoenfeld; Hsieh and Lavori 2000),^41^ cross-checked in the Dutch cohort against ‘powerSurvEpi’^42^ and ‘survSNP’^43^ (**Extended Methods**). *P*-values for the main Cox analyses were corrected for the False Discovery Rate (FDR; Benjamini-Hochberg) within each cohort; a two-sided *FDR* < 0.05 was considered significant.

#### Mediation analyses

We examined whether MMSE or ADL functioning (BI/Katz) mediated the association between plasma NfL and mortality using a regression-based g-formula implemented in the R package ‘CMAverse’^44^ (v.0.1.0), with NfL modelled as a continuous exposure. A linear model was fitted for each mediator and a Cox proportional-hazards model for mortality, both adjusted for age and sex. The decomposition assumed no exposure-mediator interaction, a specification the data supported, as all NfL by mediator interaction terms were non-significant (all *P*>0.100; **Extended Data Table 10**). Effects were estimated for a cohort-specific 25th-to-75th NfL percentile contrast. The natural direct and indirect effects, the total effect, and the proportion mediated were estimated by counterfactual imputation, with 95%CIs from 1,000 bootstrap resamples (percentile method; seed 50245); mediation was inferred when the 95%CI of the total natural indirect effect (hazard-ratio scale) excluded 1.

#### Nested model comparison

To assess the incremental prognostic value of plasma biomarkers, we compared nested Cox proportional-hazards models against an age- and sex-adjusted reference, sequentially adding clinical predictors (MMSE, BI/Katz), individual biomarkers, and their combinations. Model fit was compared using the Akaike Information Criterion (AIC; lower indicates a better fit-to-complexity trade-off) and Likelihood Ratio Tests (LRT).

#### Restricted Mean Survival Time

To express the prognostic impact of NfL in absolute terms (years of survival), we fitted flexible parametric survival models for Restricted Mean Survival Time (RMST)^45^ using the R package ‘rstpm2’.^46^ (v.1.7.1). RMST is the average expected survival up to a horizon (τ). For each cohort, a reference profile fixed age, sex, and ADL functioning (BI/Katz index) at their cohort means and NfL at its median; deviations from this reference give the difference in expected survival attributable to NfL.

RMST was evaluated at fixed time horizons of τ = 1, 2, and 3 years in the two centenarian cohorts and τ = 2, 4, and 6 years in the nonagenarian cohort. From the same models we also estimated the covariate-adjusted probability of surviving to each horizon (the survival function S(τ)) at the reference profile, with NfL set to its median, the 90th percentile, 100 pg/mL, 200 pg/mL, and the cohort maximum; 95% confidence intervals were obtained by the delta method, and model-based survival probabilities were verified against Kaplan-Meier estimates at the mean covariate profile. Analyses and visualizations used R (version 4.5.2) and Stata 16.0.

#### Missing test score imputation

For the Dutch centenarian cohort, missing scores on the MMSE and ADL functioning (BI) caused by sensory, motor impairments or fatigue were imputed using Multiple Imputation by Chained Equations (MICE) v3.13.0.^36^ Predictive mean matching was selected for its ability to handle non-normal data distributions, and 74 iterations were performed,^47^ consistent with previous publications of the same cohort (**Extended Methods; Extended Data Table 9**).^48,49^

## AUTHOR CONTRIBUTIONS

Simoa plasma biomarker concentration measurements were performed by M.G. and C.E.T. from the Neurochemistry lab at Amsterdam UMC, location VUmc. Neuropsychological assessment and collection of data for the Dutch centenarian (100-plus Study) were performed by M.J.I.G., M.E.H., D.A.H.D. and L.M.C.L. Data collection for the Danish cohorts was supervised by K.C. and M.T. Data analyses were performed by L.M.C.L. and M.T., supported by S.A.M.S., M.W.H., M.H., N.T., C.E.T., K.C. and H.H. Underlying data was verified by L.M.C.L., S.A.M.S., C.E.T. and H.H. The research was supervised by M.T., S.A.M.S., C.E.T. and H.H. Design of the research involved L.M.C.L., M.T., S.A.M.S., C.E.T. and H.H. The 100-plus Study was initiated by H.H. Design of the neurocognitive testing procedures of the 100-plus Study was supported by S.A.M.S.

## ACKNOWLEDGEMENTS

We extend our gratitude to the participants and their family members from both the Dutch and Danish cohorts for their invaluable participation. We furthermore acknowledge the research teams in both The Netherlands and Denmark for their invaluable contributions to data collection and study coordination throughout the years.

The Dutch 100-plus Study was funded by BrightFocus A2021031S, VUmc foundation, and Surf Sara (Surf 2022.031). H.H. is supported by ABOARD, a public-private partnership receiving funding from ZonMw (73305095007) and Health~Holland, Topsector Life Sciences & Health (PPP-allowance; LSHM20106), the Hans und Ilse Breuer Stiftung (2020), the HorstingStuit Foundation (2018), and the Dutch Research Council (Aspasia premie 015.016.059). The Danish studies were supported by grants from the Danish National Research Foundation and National Institute on Aging (Grant NIA P01-AG08761) and the VELUX Foundation. The funders were not involved in the study design, data collection, analysis, interpretation, or the writing of this report.

## COMPETING INTERESTS

The authors declare that they have no competing interests.

## DATA AVAILABILITY STATEMENT

The data that support the findings of this study are not publicly available due to privacy and ethical restrictions, as they contain information that could compromise participant confidentiality. Data are, however, available upon reasonable request and subject to a data sharing agreement. Inquiries can be directed to the corresponding author.

## CODE AVAILABILITY STATEMENT

The custom R code used for the statistical analyses in this study is available from the corresponding author upon reasonable request.

## Abbreviation

AD: Alzheimer disease
ADL: Activities of Daily Living
AIC: Akaike Information Criterion
Aβ: Amyloid-beta
Aβ_40_: Amyloid-beta 1-40
Aβ_42_: Amyloid-beta 1-42
BI: Barthel Index
CI: Confidence interval
FDR: False Discovery Rate
GFAP: Glial fibrillary acidic protein
HR: Hazard ratio
IQR: Interquartile range
LRT: Likelihood Ratio Test
MFP: Multivariable fractional polynomial
MICE: Multiple Imputation by Chained Equations
MMSE: Mini-Mental State Examination
N: Sample size (number of participants)
NfL: Neurofilament light
NFT: Neurofibrillary tau tangles
pTau-181: Phosphorylated tau at threonine 181
RMST: Restricted Mean Survival Time
SD: Standard deviation
Simoa: Single Molecule Array
STROBE: Strengthening the Reporting of Observational Studies in Epidemiology
τ: Time horizon tau
*z*: Z-standardized score

## Notes

### Competing Interest Statement

The authors have declared no competing interest.

### Author Declarations

The Medical Ethics Committee of Amsterdam UMC, Vrije Universiteit Amsterdam (Amsterdam, The Netherlands) gave ethical approval for this work (registration 2016.440; Dutch 100-plus Study). The Regional Committees on Health Research Ethics for Southern Denmark gave ethical approval for this work (approval nos. S-20140099 and VF20040240; Danish 1915 West Birth Cohort and 1905 Birth Cohort), with registration at the Danish Data Protection Agency (nos. 2016-41-4552 and 2020-522-0256). The Ethics Committee of the Medical Faculty of the University of Tuebingen (Germany) gave ethical approval for the biomarker analyses of the Danish cohorts (project nos. 837/2017BO2 and 838/2017BO2). All participants or their legal proxies provided written informed consent.

