## Supplementary Material (Extended Data Methods, Tables and Figures) for "Plasma neurofilament light links mortality to neuroaxonal injury beyond clinical decline and Alzheimer disease plasma biomarkers: a multi-cohort study of oldest old individuals"

**TABLE OF CONTENTS**

**Extended Data Methods**

**1.1 Study design**

**1.2 Blood-based biomarker evaluation**

1.2.1 *Blood collection*
1.2.2. *Simoa plasma biomarker measurements*

**1.3 Clinical test data**

1.3.1. *Cognitive function assessment*
1.3.2. *ADL physical functioning assessment*
1.3.3. *Missing test score imputation*

**1.4 Statistical analyses**

1.4.1. *Cox proportional hazards regression*
1.4.2. *Mediation analyses*
1.4.3. *Restricted Mean Survival Time (RMST)*
1.4.4. *Multivariable fractional polynomial (MFP) sensitivity analyses*
1.4.5. *A posteriori statistical power*

**Extended Data Tables**

*Extended Data Table 1. Tertile cut-points and distributional statistics of biomarkers*

*Extended Data Table 2. A posteriori statistical power analyses*

*Extended Data Table 3. Multivariable fractional polynomial analysis of all biomarkers and clinical predictors*

*Extended Data Table 4.	NfL attenuation by clinical functioning measures*

*Extended Data Table 5.	Mediation of the plasma NfL association with mortality through cognition (MMSE) and ADL physical functioning (BI/Katz)*

*Extended Data Table 6. Continuous-exposure mediation of each plasma biomarker on mortality, through cognition (MMSE) and physical function (Barthel ADL), Dutch centenarian cohort*

*Extended Data Table 7.	Nested models of all-cause mortality*

*Extended Data Table 8. Predicted restricted mean survival time (RMST) and covariate-adjusted survival probability
 as a function of plasma NfL concentration, by cohort and survival time horizon*

*Extended Data Table 9. Pre- and post-imputation descriptive statistics for MMSE and Barthel Index in Dutch
 centenarians*

*Extended Data Table 10.	Interaction terms between plasma biomarkers and clinical function scores each estimated in a separate multivariable Cox model for all-cause mortality*

**Extended Data Figures**

*Extended Data Figure 1. Kaplan-Meier survival curves for plasma biomarkers (except NfL) in Dutch
 centenarians*

*Extended Data Figure 2.	Forest plots of hazard ratios for all-cause mortality associated with demographics, plasma biomarkers and multi-item scales for cognitive and ADL physical functioning*

*Extended Data Figure 3.	Distributions of plasma biomarkers, MMSE scores and ADL scores*

**Extended Data References**

Extended Data Methods

1.1 Study design

**Dutch centenarians:** The 100-plus Study is an ongoing longitudinal prospective cohort study comprising centenarians who self-report to be cognitively healthy at baseline, as confirmed by proxy. Detailed descriptions of the cohort’s collection procedures and characteristics have been published previously.^1^ All centenarians were born between 1909 and 1923 and had no clinical diagnosis of dementia or signs of neurological disorders at the time of inclusion. Baseline home visits involved collecting comprehensive lifetime histories, including educational backgrounds, assessing cognitive performance and physical functioning, and collecting blood samples. The cognitive testing (MMSE) and the physical function assessment (BI) were conducted on the same day as blood collection for the majority of centenarians (n=217, 85%); for the remaining centenarians (n=38, 15%), there was a median interval of 3.1 months (IQR 1.7–5.3) between the clinical and blood assessments.

**Danish centenarians:** The Danish 1915 West birth cohort study included individuals who were born in 1915, alive on their 100th birthday, and living in the Western part of Denmark. Centenarians were assessed in their own homes or in care facilities. From the initial 303 centenarians, 238 participated in a health survey. In-person interviews were conducted for 185 participants, and 131 provided blood samples. With four additional centenarians born in 1914 from a 2014 pilot study, 135 had blood samples available, of whom 128 also had complete clinical data and were included in the present analyses.

**Danish nonagenarians:** The Danish 1905 birth cohort included individuals born in 1905 who were alive on August 1, 1998. Out of the 3,725 eligible nonagenarians, 2,262 participated in a comprehensive health survey. For the 231 participants living in Funen, whole-blood samples were collected for 180 participants by a physician shortly after the in-person interviews and of these, 176 also had complete clinical data and were included in the present analyses.

1.2 Blood-based biomarker evaluation

1.2.1 Blood collection

**Dutch centenarians:** The higher age at blood collection observed in decedents reflects the fact that plasma biomarker sampling was introduced after cohort enrolment had begun. Blood samples were collected via venipuncture into 6 mL EDTA K2 tubes and transported at room temperature during transit to the lab. Sample preparation involved centrifugation at 1800xg for 10 minutes, followed by storage in 0.5 mL aliquots at -80°C. Before assaying, samples were thawed at room temperature and centrifuged at 10,000xg and 4°C for five minutes.

**Danish centenarians & nonagenarians:** Whole-blood samples were collected in 10-ml EDTA tubes. To ensure sample integrity, the blood was centrifuged at 1,000xg for 15 minutes on the same day that the donation took place. Plasma was subsequently aliquoted and directly stored at -80°C until biomarker analysis.

1.2.2 Simoa plasma biomarker measurements

**Dutch centenarians**: All assays were performed at the Neurochemistry Laboratory of Amsterdam UMC. Plasma pTau-181 was analyzed using the Simoa pTau-181 Advantage V2.0 Kit. Concentrations of Aβ_40_, Aβ_42_, NfL, and GFAP were analyzed simultaneously with the Neurology 4-Plex-E Advantage (N4PE) Kit on a Simoa HD-X analyzer (Quanterix, Billerica, MA, USA). All plasma samples were labeled with a random code and randomized across assay batches. All measured biomarker concentrations were above the lower limit of quantification (LLOQ).

For pTau-181, intra-assay and inter-assay coefficient of variation (CV) percentages were 6.1% and 9.2% for low quality control (QC), and 11.4% for high QC. Quality control included two control samples per biomarker on each plate, achieving a CV% below 20%. To preserve data integrity, three participants with three freeze-thaw cycles were excluded from GFAP analyses.

**Danish centenarians & nonagenarians:** Plasma NfL concentrations were determined using the Simoa NF-Light Advantage kit (Quanterix) strictly adhering to the manufacturer's instructions. Before measurement, all plasma samples were centrifuged for 5 min at 10,000xg and 4°C. Samples were diluted 1:4 with Simoa NF-Light sample diluent. All measurements were performed on a Simoa HD-1 or HD-X Analyzer. All samples were measured in duplicate, and laboratory personnel were blinded to all clinical and demographic data; quality control was maintained through internal reference samples on every plate.

1.3 Clinical test data

1.3.1 Cognitive function assessment

Across all cohorts, given the high prevalence of sensory impairments in this age group, MMSE items that participants were unable to complete due to visual or hearing loss were excluded, and the total score was rescaled to a maximum of 30 to maintain comparability across participants.

**Dutch centenarians**: Global cognitive functioning was assessed using a modified version of the Mini-Mental State Examination (MMSE)^2^^,^^3^ (scores range from 0 to 30, with higher scores indicating better cognitive performance). The MMSE is a widely used screening instrument for evaluating general cognitive status and encompasses multiple cognitive domains, including orientation to time and place, short-term memory (immediate and delayed recall), attention and concentration (e.g., serial sevens, spelling), language (naming, repetition, comprehension, reading, and writing), and visuospatial skills (e.g., figure copying).

**Danish centenarians & nonagenarians:** The health surveys similarly included cognitive performance assessments using the MMSE, ensuring comparability of general cognitive status across the cohorts.

1.3.2 ADL physical functioning assessment

**Dutch centenarians**: Functional status was assessed using the Barthel Index (BI)^4^ of Activities of Daily Living (ADL; scores range from 0 to 20, with higher scores indicating greater independence). The Barthel Index evaluates a person’s ability to perform ten basic self-care and mobility activities, including feeding, bathing, grooming, dressing, bowel and bladder continence, toilet use, transfers, mobility, and stair climbing. Each activity is scored based on the level of assistance required, providing a cumulative measure of functional independence in daily life. Assessments were conducted in participants' homes or care facilities by trained research personnel

**Danish centenarians & nonagenarians:** ADL was assessed by self-report of 11 physical tasks. Of these, the five items assessing bathing, dressing, toileting, transfer, and feeding were used to derive the ADL disability score, which is based on the standardized Katz ADL index.^5^ Because the Katz index has a narrow discrete range (0–5), cohort-specific tertile boundaries differed between the two Danish cohorts, reflecting their distinct functional distributions. In the nonagenarian cohort, where ADL functioning was skewed toward independence, tertiles were defined as T1 (lower functioning; scores 0–3), T2 (middle; score 4), and T3 (higher functioning; score 5). In the centenarian cohort, where greater functional impairment shifted the distribution downward, tertiles were defined as T1 (lower functioning; scores 0–2), T2 (middle; score 3), and T3 (higher functioning; scores 4–5). This approach ensures that tertile membership reflects relative functioning within each cohort rather than imposing uniform cut-points that would misrepresent the proportion of individuals at each functional level. Assessments were conducted by trained lay personnel (1905 birth cohort) and by a geriatrician and a nurse (1915 West birth cohort).

1.3.3 Missing test score imputation

**Dutch centenarians**: For the MICE procedure,^6^^,^^7^ we utilized data from 472 centenarians included in the study at the time of analysis. This included age at cognitive assessment, sex, all available neuropsychological test scores, Geriatric Depression Scale (GDS) scores,^8^ Barthel Index (BI) scores for activities of daily living (ADL) functioning,^4^ vision and hearing abilities, educational attainment, Informant Questionnaire on Cognitive Decline in the Elderly (IQCODE) scores,^9^ test time limit exceedances and qualitative evaluations of cognitive status, understanding, and motivation to complete the tests. Missing MMSE items were individually imputed, accounting for sensory function variability among centenarians. Predictive mean matching was selected for its ability to handle non-normal data distributions, and 74 iterations were performed, consistent with a previous publication of the same cohort.^1^ Following imputation, MMSE scores were available for 249 centenarians. Centenarians with six or more missing responses on the MMSE (n=6, 2%) were excluded from the analyses.

**Danish centenarians & nonagenarians:** No imputation was applied.

1.4 Statistical analyses

1.4.1. Cox proportional hazards regression

To assess the robustness of the association of each predictor with all-cause mortality, we fit a series of progressively adjusted Cox regression models. In Models 1 to 5, every predictor was entered in its own separate model, so that each hazard ratio reflects the effect of a single predictor adjusted only for the covariates listed in the column header and not for the other biomarkers. The models were specified as follows: (Model 1) adjusted for age and sex; (Model 2) adjusted for age, sex, and MMSE score; (Model 3) adjusted for age, sex, and ADL functioning; (Model 4) adjusted for age, sex, MMSE, and ADL functioning; (Model 5) adjusted for age, sex, MMSE, ADL functioning, and NfL.

In contrast, Model 6 was a single simultaneous model in which all biomarkers (Aβ_42/40_ ratio, Aβ_40_, Aβ_42_, pTau-181, pTau-181/Aβ_42_ ratio, NfL, and GFAP) were entered together in one model, alongside age, sex, MMSE, and ADL functioning. Model 6 therefore estimates the independent contribution of each biomarker while mutually adjusting for all other biomarkers. Model 6 was fitted for the Dutch centenarian cohort only.

For the predictor rows NfL, MMSE, and ADL functioning, cells in which a predictor would need to adjust for itself are left blank, as the model reduces to a lower nested specification already shown. Formal testing of the proportional-hazards assumption via Schoenfeld residuals revealed that no predictors violated proportionality, except for plasma GFAP in the Dutch cohort (χ² = 6.97, *P*=0.008). Given that GFAP showed no significant association with mortality in any model, this violation was considered inconsequential for the primary analyses.

To test whether the prognostic value of each plasma biomarker depended on cognitive or physical functioning, we additionally fitted, for each biomarker, a separate Cox model containing the biomarker, one clinical score (MMSE or ADL functioning), their interaction, and age and sex. Only the interaction terms are reported (**Extended Data Table 10**); no interaction was statistically significant.

To compare the incremental prognostic value of the biomarkers and clinical scores, we fitted a set of nested Cox models within each cohort, each adding one or more predictors to a demographic reference model (age and sex). Each candidate model was compared with the reference using a likelihood ratio test, and model fit was summarized with the Akaike information criterion (AIC); a negative ΔAIC indicates improved fit relative to the reference (**Extended Data Table 7**). To keep the likelihood ratio tests and AIC values comparable, all models within a cohort were fitted on the same participants with complete data on every variable.

1.4.2. Mediation analyses

We examined whether cognitive performance (MMSE) or physical functioning (ADL; Barthel Index in the Dutch cohort, Katz Index in the Danish cohorts) mediated the association between plasma NfL and all-cause mortality, using the regression-based ("rb") approach of the natural effects framework implemented in the R package "CMAverse" (v.0.1.0).^10^ NfL was modelled as a continuous exposure throughout; it was never dichotomized. For each mediator we fitted two regressions adjusted for age and sex: a linear model for the mediator given NfL and covariates (the exposure-to-mediator path), and a Cox proportional-hazards model for mortality given NfL, the mediator, and covariates (the mediator-to-outcome and direct paths). The decomposition assumed no exposure-mediator interaction; this specification was supported by the data, as all NfL-by-mediator interaction terms were non-significant (all *P*>0.100; **Extended Data Table 10**).

Effects were estimated for a cohort-specific contrast between the 25th and 75th percentiles of the observed NfL distribution, so that each estimate expresses the effect of an interquartile increase in NfL. From the fitted regressions, CMAverse derived the pure and total natural direct effects, the pure and total natural indirect effects, the total effect, and the proportion mediated by direct counterfactual imputation, expressing all effects on the hazard-ratio scale. The use of natural effect models for mediation with a time-to-event outcome follows the approach evaluated by Lange et al. and Burgos Ochoa et al.^11^^,^^12^ Uncertainty was quantified by nonparametric bootstrap with 1,000 resamples, from which 95% confidence intervals were obtained by the percentile method (2.5th and 97.5th quantiles of the bootstrap distribution); a fixed seed (50245) was set for reproducibility. Mediation through a given mediator was inferred when the 95% confidence interval of the total natural indirect effect (hazard-ratio scale) excluded 1. The same procedure was applied to each plasma biomarker as a continuous exposure in the Dutch centenarian cohort (**Extended Data Table 6**); for biomarkers other than NfL the total effect was null, so the proportion mediated is undefined and is not reported.

1.4.3. Restricted Mean Survival Time (RMST)

Flexible parametric models were specified with restricted cubic splines (4 degrees of freedom) for the baseline hazard. NfL entered as a continuous unstandardized variable (pg/mL). Predicted RMST curves were generated across each cohort's observed NfL range, with 95% confidence intervals obtained via the delta method. Effect modification between NfL and ADL functioning was tested by adding interaction terms and evaluated by likelihood ratio test. Time horizons were 1, 2, and 3 years for the two centenarian cohorts (Dutch and Danish), and 2, 4, and 6 years for the Danish nonagenarians. The change in the slope of the RMST curve with respect to NfL concentration was compared across time horizons to quantify how the prognostic signal changes over longer survival periods.

1.4.4. Multivariable fractional polynomial (MFP) sensitivity analyses

To test the robustness of our attenuation analyses to non-linear relationships, we performed MFP analyses on NfL, MMSE, and ADL functioning simultaneously in all three cohorts, adjusting for age and sex. In the Dutch centenarian cohort, where the full plasma panel was available, the same procedure was additionally applied to each plasma biomarker, adjusting for age, sex, MMSE, and ADL functioning (with MMSE and the Barthel Index also entered as fractional-polynomial terms). Full results and partial-effect plots are available in **Extended Data Table 3**.

As a targeted check on the linear specification of ADL functioning in the attenuation models, we refitted the NfL models entering ADL in its functional form and compared the NfL hazard ratio and its attenuation against the linear specification. In the Dutch centenarian cohort, where MFP selected a cubic form for the Barthel Index, the NfL hazard ratio was essentially unchanged whether ADL was entered linearly or as a cubic. In the two Danish cohorts MFP selected a linear form for the Katz index, so the linear specification is already correct; as a conservative bound we re-ran the same comparison forcing the Dutch-derived cubic transformation, and the NfL hazard ratio again changed only marginally and remained significant. Full results across the three cohorts are reported in **Extended Data Table 4**.

Multivariable fractional polynomial (MFP) modelling tested non-linearity for NfL, MMSE, and ADL. MMSE was linear (power = 1) in all cohorts; NfL was linear in both centenarian cohorts and log-transformed (power = 0) in the nonagenarians; ADL was linear in the Danish cohorts but cubic (power = 3) in the Dutch. NfL remained significant under the MFP-selected forms in all three cohorts (Dutch HR 1.24, 95%CI 1.09 to 1.40, *P*=0.001; Danish centenarians HR 1.38, 95%CI 1.17 to 1.64, *P*<0.001; Danish nonagenarians HR 1.58, 95%CI 1.27 to 1.97, *P*<0.001), confirming its independent value beyond MMSE and ADL is not a linear-specification artefact (**Extended Data Table 3**). In the Dutch cohort, the cubic-ADL model attenuated NfL by 17% (HR 1.24 from Model 1 HR 1.29 on the log-HR scale), essentially identical to the 16% under linear ADL (**Extended Data Table 4**).

1.4.5. A posteriori statistical power

A posteriori statistical power was summarized for each cohort as the minimum hazard ratio (HR) per 1-SD increase in a predictor that the design could detect at 80% and 90% power under a two-sided α of 0.05, conditional on the number of observed deaths. Three estimators were used. The first was an analytical events-based formula (Schoenfeld; Hsieh and Lavori 2000),^13^ HR_min_ = exp[(z_1-α/2_ + z_1-β_) / √(d(1-ρ²))], where d is the number of deaths and ρ² is the squared multiple correlation between the predictor and the age and sex covariates. The second was numerical inversion of powerSurvEpi^14^::powerEpiCont, solved by binary search on the effect size to reach the target power. The third was a per-SD conversion of survSNP^15^::survSNP.power.table in which the per-allele HR was inverted by binary search and converted to a per-SD HR via HR_per-SD_ = HR_per-allele_^√(0.5/(1-ρ²))^.

In the Dutch centenarian cohort, follow-up was incomplete (91% events). For NfL and the Barthel Index, 233 deaths were observed among 255 participants; for MMSE, 228 deaths among 249 participants with complete model covariates. Across these predictors the analytical, powerSurvEpi and survSNP estimators converged to within approximately 0.01. In both Danish cohorts follow-up was complete (100% events); because powerSurvEpi and survSNP require a non-zero censoring fraction (the variance of the event indicator is zero when no participant is censored), both returned undefined values, and the analytical events-based formula, which depends only on the number of deaths, was used.

Because the AD-specific plasma biomarkers were measured only in the Dutch centenarian cohort, this analysis is inferential for the AD-biomarker null findings in that cohort: the AD-specific biomarkers (Aβ_40_, Aβ_42_, the Aβ_42/40_ ratio, pTau-181, the pTau-181/Aβ_42_ ratio) and GFAP had observed effects at or below the 1.21 minimum detectable HR at 80% power (**Table 2**). In the two Danish cohorts, where only NfL was measured and all predictors were associated with mortality, the analysis is reported as a descriptive index of cohort sensitivity. Per-cohort minimum detectable HRs are reported in **Extended Data Table 2**.

**Extended Data Tables**

| **Extended Data Table 1. Tertile cut-points and distributional statistics of biomarkers and clinical measures** | | | |
| --- | --- | --- | --- |
|  | **Tertile 1,  Upper limit,  median (min, max)** | **Tertile 2, Upper limit,  median (min, max)** | **Tertile 3, Upper limit,  median (min, max)** |
| **Dutch centenarians** |  |  |  |
| Aβ_42/40_ ratio | 0.050, 0.045 (0.017, 0.050) | 0.056, 0.053 (0.050, 0.056) | 0.087, 0.062 (0.056, 0.087) |
| Aβ_40_ | 139.2, 114.9 (68.6, 139.2) | 176.4, 155.2 (139.9, 176.4) | 299.2, 209.3 (176.5, 299.2) |
| Aβ_42_ | 6.85, 5.93 (2.32, 6.85) | 9.26, 8.07 (6.87, 9.26) | 17.66, 10.83 (9.28, 17.66) |
| pTau-181 | 3.49, 2.76 (1.46, 3.49) | 4.85, 4.05 (3.49, 4.85) | 12.75, 6.42 (4.88, 12.75) |
| pTau-181/Aβ_42_ ratio | 0.431, 0.320 (0.161, 0.431) | 0.653, 0.534 (0.434, 0.653) | 2.459, 0.849 (0.655, 2.459) |
| NfL | 40.8, 32.4 (11.9, 40.8) | 63.6, 50.5 (41.0, 63.6) | 378.6, 88.8 (63.7, 378.6) |
| GFAP | 207.0, 169.2 (88.0, 207.0) | 293.2, 251.8 (207.4, 293.2) | 1085.1, 361.0 (293.7, 1085.1) |
| MMSE score | 24.0, 21.0 (12.0, 24.0) | 27.0, 26.0 (24.1, 27.0) | 30.0, 28.8 (27.2, 30.0) |
| BI score | 13.5, 10.0 (0.0, 13.5) | 17.0, 16.0 (14.0, 17.0) | 20.0, 19.0 (17.0, 20.0) |
| **Danish centenarians** |  |  |  |
| NfL | 42.0, 34.0 (22.0, 42.0) | 59.8, 49.4 (42.1, 59.8) | 460.9, 88.8 (60.0, 460.9) |
| MMSE score | 17.0, 13.0 (5.0, 17.0) | 23.0, 21.0 (18.0, 23.0) | 29.0, 26.0 (24.0, 29.0) |
| Katz score | 2.0, 1.0 (0.0, 2.0) | 3.0, 3.0 (3.0, 3.0) | 5.0, 5.0 (4.0, 5.0) |
| **Danish nonagenarians** |  |  |  |
| NfL | 29.6, 22.9 (13.9, 29.6) | 45.3, 34.8 (29.8, 45.3) | 384.7, 62.9 (45.5 , 384.7) |
| MMSE score | 20.0, 17.0 (7.0, 20.0) | 25.0, 24.0 (21.0, 25.0) | 30.0, 27.0 (26.0, 30.0) |
| Katz score | 3.0, 2.0 (0.0, 3.0) | 4.0, 4.0 (4.0, 4.0) | 5.0, 5.0 (5.0, 5.0) |
| *Abbreviations*: Aβ, amyloid-β; BI, Barthel Index; GFAP, glial fibrillary acidic protein; MMSE, Mini-Mental State Examination; NfL, neurofilament light chain; pTau-181, phosphorylated tau at threonine 181. Tertile cut-points were defined using the 33rd and 66th percentiles of the baseline distribution within each cohort. All plasma biomarkers are expressed in pg/mL, except for the unitless ratios. Sample sizes: Dutch centenarians (N=255 for plasma biomarkers and BI; N=249 for MMSE), Danish centenarians (N=128), and Danish nonagenarians (N=176). | | | |

| **Extended Data Table 2. A posteriori statistical power analyses** | | | | | |
| --- | --- | --- | --- | --- | --- |
| **Predictor** | **Observed HR per SD (95% CI)** | ***P*** | **ρ²  (age, sex)** | **Min. detectable HR per SD, 80% power** | **Min. detectable HR per SD, 90% power** |
| **Dutch centenarians** | | | | | |
| NfL (positive control) | 1.29 (1.15 to 1.45) | **<0.001** | n.a. | 1.21 | 1.25 |
| Aβ_40_ | 1.09 (0.95 to 1.26) | 0.195 | 0.055 | 1.21 | 1.24 |
| Aβ_42_ | 1.08 (0.94 to 1.24) | 0.252 | 0.034 | 1.20 | 1.24 |
| Aβ_42/40_ ratio | 1.00 (0.87 to 1.15) | 0.998 | 0.007 | 1.20 | 1.24 |
| pTau-181 | 1.02 (0.88 to 1.18) | 0.840 | 0.069 | 1.21 | 1.25 |
| pTau-181/Aβ_42_ ratio | 0.93 (0.82 to 1.05) | 0.238 | 0.012 | 1.20 | 1.24 |
| GFAP | 1.11 (0.97 to 1.27) | 0.135 | 0.071 | 1.21 | 1.25 |
| MMSE | 0.84 (0.73 to 0.96) | **0.009** | 0.032 | 1.21 | 1.24 |
| Barthel Index | 0.73 (0.63 to 0.83) | **<0.001** | 0.142 | 1.22 | 1.26 |
| **Danish centenarians** |  |  |  |  |  |
| NfL | 1.50 (1.271 to 1.770) | **<0.001** | 0.006 | 1.28 | 1.33 |
| MMSE | 0.67 (0.555 to 0.806) | **<0.001** | 0.011 | 1.28 | 1.33 |
| Katz Index | 0.73 (0.610 to 0.873) | **<0.001** | 0.049 | 1.29 | 1.34 |
| **Danish nonagenarians** |  |  |  |  |  |
| NfL | 1.22 (1.101 to 1.361) | **<0.001** | 0.012 | 1.24 | 1.28 |
| MMSE | 0.79 (0.678 to 0.916) | **0.002** | 0.001 | 1.25 | 1.28 |
| Katz Index | 0.72 (0.607 to 0.842) | **<0.001** | 0.008 | 1.24 | 1.28 |
| *Abbreviations*: Aβ, amyloid-β; GFAP, glial fibrillary acidic protein; HR, hazard ratio; MMSE, Mini-Mental State Examination; n.a., not applicable; NfL, neurofilament light chain; pTau-181, phosphorylated tau at threonine 181; SD, standard deviation. Values are the minimum HR per 1-SD increase detectable at 80% and 90% power under a two-sided α of 0.05, given the observed number of deaths; ρ² is the squared multiple correlation between the predictor and the age and sex covariates. Full methods, including the estimator formula, the per-cohort event counts, and the treatment of the AD-specific biomarkers, are given in **Supplementary Methods 1.4.5.** | | | | | |

| **Extended Data Table 3. Multivariable fractional polynomial analysis of all biomarkers and clinical predictors** | | | |
| --- | --- | --- | --- |
|  | **Selected power** | **HR (95% CI)** | ***P*-value** |
| **Dutch centenarians** |  |  |  |
| Aβ_42/40_ ratio | 1 (linear) | 0.97 (0.84, 1.12) | 0.701 |
| Aβ_40_ | 1 (linear) | 1.11 (0.97, 1.28) | 0.146 |
| Aβ_42_ | 1 (linear) | 1.08 (0.93, 1.24) | 0.315 |
| pTau-181 | -1 (reciprocal)^a^ | 1.10 (1.03, 1.18) | **0.005** |
| pTau-181/Aβ_42_ ratio | 1 (linear) | 0.93 (0.82, 1.06) | 0.269 |
| GFAP | 1 (linear) | 1.08 (0.94, 1.24) | 0.274 |
| NfL | 1 (linear) | 1.24 (1.09, 1.40) | **0.001** |
| MMSE score (NfL model) | 1 (linear) | 0.97 (0.84, 1.12) | 0.678 |
| BI score (NfL model) | 3 (cubic)^b^ | n.a.^b^ | **<0.001** |
| **Danish centenarians** |  |  |  |
| NfL | 1 (linear) | 1.38 (1.17, 1.64) | **<0.001** |
| MMSE score | 1 (linear) | 0.77 (0.62, 0.95) | **0.014** |
| Katz score | 1 (linear) | 0.82 (0.66, 1.00) | 0.053 |
| **Danish nonagenarians** |  |  |  |
| NfL | 0 (log) | 1.58 (1.27, 1.97) | **<0.001** |
| MMSE score | 1 (linear) | 0.91 (0.77, 1.08) | 0.273 |
| Katz score | 1 (linear) | 0.84 (0.70, 1.01) | 0.057 |
| *Abbreviations*: Aβ, amyloid-β; BI, Barthel Index; CI, confidence interval; GFAP, glial fibrillary acidic protein; HR, hazard ratio; MFP, multivariable fractional polynomial; MMSE, Mini-Mental State Examination; n.a., not applicable; NfL, neurofilament light chain; pTau-181, phosphorylated tau at threonine 181; SD, standard deviation. In Dutch centenarians, each biomarker was tested in a separate model together with MMSE and the Barthel Index, all entered as fractional-polynomial terms, adjusted for age and sex; the MMSE and Barthel rows are from the NfL model. In the Danish cohorts, NfL, MMSE, and Katz were entered as fractional-polynomial terms and tested simultaneously, adjusted for age and sex. HRs for linear forms are per 1-SD increase. ^a^HR on the transformed scale, not comparable to per-SD HRs. This reciprocal form for pTau-181 was selected by the data-driven MFP search and was not significant under the pre-specified linear model (**Table 2**); its P-value is therefore not adjusted for the functional-form selection and is anti-conservative, and the result was not corroborated by the mediation, nested-model, or interaction analyses. It should be interpreted as an artefact of flexible modelling rather than as evidence of a pTau-181 association with mortality. ^b^Cubic form; no single per-SD HR (steepest at low Barthel scores). Sample sizes: Dutch centenarians N=249 (GFAP N=246), Danish centenarians N=128, and Danish nonagenarians N=176. | | | |

| **Extended Data Table 4. NfL attenuation by clinical functioning measures** | | | |
| --- | --- | --- | --- |
| **Model** | **NfL HR per SD (95% CI)** | ***P*** | **Attenuation vs M1 (%)** |
| **Dutch centenarians** | | | |
| M1: NfL + age, sex (reference) | 1.29 (1.14, 1.45) | **<0.001** | reference |
| M2: + MMSE | 1.26 (1.12, 1.43) | **<0.001** | ~9 |
| M3: + ADL (linear) | 1.24 (1.09, 1.41) | **<0.001** | ~14 |
| M3: + ADL (cubic, MFP) | 1.24 (1.09, 1.40) | **<0.001** | ~16 |
| M4: + MMSE + ADL (linear) | 1.24 (1.09, 1.41) | **0.001** | ~17 |
| M4: + MMSE + ADL (cubic, MFP) | 1.24 (1.09, 1.40) | **0.001** | ~17 |
| **Danish centenarians** | | | |
| M1: NfL + age, sex (reference) | 1.50 (1.27, 1.77) | **<0.001** | reference |
| M2: + MMSE | 1.38 (1.16, 1.64) | **<0.001** | ~20 |
| M3: + ADL (linear) | 1.45 (1.23, 1.71) | **<0.001** | ~8 |
| M4: + MMSE + ADL (linear) | 1.38 (1.17, 1.64) | **<0.001** | ~20 |
| **Danish nonagenarians** | | | |
| M1: NfL + age, sex (reference) | 1.22 (1.10, 1.36) | **<0.001** | reference |
| M2: + MMSE | 1.22 (1.09, 1.37) | **<0.001** | ~1 |
| M3: + ADL (linear) | 1.21 (1.08, 1.36) | **0.001** | ~4 |
| M4: + MMSE + ADL (linear) | 1.21 (1.08, 1.36) | **0.001** | ~5 |
| *Abbreviations*: ADL, activities of daily living; HR, hazard ratio; MFP, multivariable fractional polynomial; MMSE, Mini-Mental State Examination; NfL, neurofilament light chain; SD, standard deviation. NfL HR per 1-SD increase, adjusted for age and sex (M1), then adding MMSE (M2), ADL (M3), and both (M4); attenuation is the percentage reduction in the NfL log-HR versus M1. ADL was entered in the MFP-selected form per cohort: Dutch centenarians (N=249, Barthel Index, cubic), Danish centenarians (N=128, Katz Index, linear), Danish nonagenarians (N=176, Katz Index, linear). In the Dutch cohort ADL is shown both linearly and in the cubic form, which give nearly identical NfL HRs. NfL remained significant throughout. | | | |

| **Extended Data Table 5. Mediation of the plasma NfL association with mortality through cognition (MMSE) and ADL physical functioning (BI/Katz)** | | | | | |
| --- | --- | --- | --- | --- | --- |
| **Mediator** | **Direct effect, PNDE HR (95% CI)** | **Indirect effect, TNIE HR (95% CI)** | **Proportion mediated, % (95% CI)** | | **Mediation** |
| **Dutch centenarians** |  |  |  | |  |
| MMSE | 1.20 (1.11, 1.48) | 1.014 (0.999, 1.045) | 7.70 (−0.70, 24.60) | | No |
| Barthel Index (ADL) | 1.18 (1.11, 1.32) | 1.038 (1.012, 1.079) | 19.50 (6.80, 35.30) | | Yes |
| **Danish centenarians** | | | | | |
| MMSE | 1.23 (1.16, 1.49) | 1.072 (1.032, 1.163) | 27.70 (12.40, 47.70) | | Yes |
| Katz Index (ADL) | 1.27 (1.20, 1.56) | 1.032 (1.004, 1.083) | 13.10 (1.40, 29.10) | | Yes |
| **Danish nonagenarians** | | | | | |
| MMSE | 1.13 (1.07, 1.58) | 1.018 (0.999, 1.068) | 13.20 (−1.70, 25.90) | | No |
| Katz Index (ADL) | 1.13 (1.06, 1.54) | 1.031 (1.003, 1.099) | 21.10 (3.30, 36.80) | | Yes |
| *Abbreviations*: ADL, activities of daily living; CI, confidence interval; HR, hazard ratio; MMSE, Mini-Mental State Examination; NfL, neurofilament light chain; PNDE, pure natural direct effect; TNIE, total natural indirect effect. NfL was modelled as a continuous exposure, each effect contrasting its 75th versus 25th percentile, adjusted for age and sex, using a regression-based g-formula with a Cox outcome and a linear mediator (1000 bootstrap resamples, percentile 95% CI). The PNDE is the part of the NfL effect not acting through the mediator and the TNIE the part acting through it; mediation is declared when the TNIE 95% CI excludes 1. | | | | | |

| **Extended Data Table 6. Continuous-exposure mediation of each plasma biomarker on mortality, through cognition (MMSE) and physical function (Barthel ADL), Dutch centenarian cohort** | | | | | | |
| --- | --- | --- | --- | --- | --- | --- |
| **Exposure (biomarker)** | **Mediator** | **N** | **Direct effect, PNDE HR (95% CI)** | **Indirect effect, TNIE HR (95% CI)** | **Proportion mediated,  % (95% CI)** | **Mediation** |
| **Dutch centenarians** |  |  |  |  |  |  |
| Aβ_42/40_ ratio | MMSE | 249 | 1.01 (0.86, 1.16) | 0.99 (0.96, 1.01) | ND | No |
| Aβ_42/40_ ratio | Barthel Index | 255 | 0.97 (0.82, 1.12) | 1.00 (0.96, 1.04) | ND | No |
| Aβ_40_ | MMSE | 249 | 1.13 (0.95, 1.40) | 0.99 (0.96, 1.02) | ND | No |
| Aβ_40_ | Barthel Index | 255 | 1.13 (0.96, 1.38) | 1.03 (0.99, 1.09) | ND | No |
| Aβ_42_ | MMSE | 249 | 1.14 (0.93, 1.39) | 0.98 (0.94, 1.01) | ND | No |
| Aβ_42_ | Barthel Index | 255 | 1.10 (0.91, 1.31) | 1.03 (0.99, 1.10) | ND | No |
| pTau-181 | MMSE | 249 | 1.02 (0.84, 1.23) | 0.99 (0.97, 1.02) | ND | No |
| pTau-181 | Barthel Index | 255 | 1.01 (0.86, 1.23) | 1.02 (0.97, 1.07) | ND | No |
| pTau-181/Aβ_42_ ratio | MMSE | 249 | 0.92 (0.81, 1.16) | 1.00 (0.98, 1.03) | ND | No |
| pTau-181/Aβ_42_ ratio | Barthel Index | 255 | 0.94 (0.82, 1.20) | 0.99 (0.95, 1.03) | ND | No |
| GFAP | MMSE | 246 | 1.09 (0.91, 1.37) | 1.02 (0.99, 1.05) | ND | No |
| GFAP | Barthel Index | 252 | 1.08 (0.93, 1.35) | 1.01 (0.96, 1.04) | ND | No |
| *Abbreviations:* CI, confidence interval; HR, hazard ratio; MMSE, Mini-Mental State Examination; ND, not defined; PNDE, pure natural direct effect; TNIE, total natural indirect effect. Each biomarker was modelled as a continuous exposure; effects contrast its 75th versus 25th percentile, computed from this cohort, adjusted for age and sex (regression-based g-formula, Cox outcome, linear mediator, 1000 bootstrap resamples, percentile CI). Mediation is declared when the TNIE 95% CI excludes 1. Proportion mediated is reported only where the overall (total) effect is non-null; for all biomarkers other than NfL the total effect was not significant, so the proportion mediated is undefined and unstable (estimates and intervals spanning hundreds of percent) and is shown as ND; the raw values are in the source CSV. All biomarkers were measured in the Dutch centenarian cohort only. Sample sizes: Dutch centenarians (N=255 for plasma biomarkers and BI; N=249 for MMSE), Danish centenarians (N=128), and Danish nonagenarians (N=176). | | | | | | |

| **Extended Data Table 7. Nested models of all-cause mortality** | | |
| --- | --- | --- |
| **Model Specification** | ***P*-value (vs Ref)** | **ΔAIC (vs Ref)** |
| *Ref: Demographic model (age and sex)* | n.a. | n.a. |
| **Dutch centenarians** |  |  |
| + Aβ_42/40_ ratio only | 1.000 | 2.00 |
| + Aβ_40_ only | 0.142 | -0.15 |
| + Aβ_42_ only | 0.214 | 0.46 |
| + pTau-181 only | 0.855 | 1.97 |
| + pTau-181/Aβ_42_ ratio only | 0.193 | 0.30 |
| + GFAP only | 0.150 | -0.07 |
| *NfL models* |  |  |
| + NfL only | **<0.001** | -10.58 |
| + NfL + MMSE | **<0.001** | -12.49 |
| + NfL + BI | **<0.001** | -23.31 |
| + NfL + MMSE + BI | **<0.001** | -21.48 |
| *Clinical models* |  |  |
| + MMSE only | **0.014** | -4.05 |
| + BI only | **<0.001** | -16.78 |
| + MMSE + BI | **<0.001** | -15.17 |
| **Danish centenarians** |  |  |
| *NfL models* |  |  |
| + NfL only | **<0.001** | -12.80 |
| + NfL + MMSE | **<0.001** | -23.01 |
| + NfL + Katz | **<0.001** | -20.67 |
| + NfL + MMSE + Katz | **<0.001** | -24.63 |
| *Clinical models* |  |  |
| + MMSE only | **<0.001** | -15.24 |
| + Katz only | **<0.001** | -9.33 |
| + MMSE + Katz | **<0.001** | -16.61 |
| **Danish nonagenarians** |  |  |
| *NfL models* |  |  |
| + NfL only | **0.002** | -7.71 |
| + NfL + MMSE | **<0.001** | -13.83 |
| + NfL + Katz | **<0.001** | -17.81 |
| + NfL + MMSE + Katz | **<0.001** | -18.50 |
| *Clinical models* |  |  |
| + MMSE only | **0.002** | -7.18 |
| + Katz only | **<0.001** | -12.32 |
| + MMSE + Katz | **<0.001** | -13.36 |
| *Abbreviations*: Aβ, amyloid-β; AIC, Akaike Information Criterion; BI, Barthel Index; GFAP, glial fibrillary acidic protein; MMSE, Mini-Mental State Examination; n.a., not applicable; NfL, neurofilament light chain; pTau-181, phosphorylated tau at threonine 181. All *P*-values and ΔAIC values are from comparisons with a reference Cox model including age and sex only. ΔAIC represents the difference in AIC between each candidate model and the reference model; negative values indicate improved fit relative to the reference. Analyses were restricted to participants with complete data on all variables: Dutch centenarians (N=246), Danish centenarians (N=128), and Danish nonagenarians (N=176). | | |

| **Extended Data Table 8. Predicted restricted mean survival time (RMST) and covariate-adjusted survival probability as a function of plasma NfL concentration, by cohort and survival time horizon** | | | | | | | | | |
| --- | --- | --- | --- | --- | --- | --- | --- | --- | --- |
| **Plasma NfL (pg/mL)** | **1 y: RMST (95% CI)** | **1 y: Δ (months)** | **1 y: S(τ), % (95% CI)** | **2 y: RMST (95% CI)** | **2 y: Δ (months)** | **2 y: S(τ), % (95% CI)** | **3 y: RMST (95% CI)** | **3 y: Δ (months)** | **3 y: S(τ), % (95% CI)** |
| **Dutch centenarians (N = 255) — horizons 1, 2, 3 years** | | | | | | | | | |
| 11.9 (min) | 0.86 (0.83, 0.90) | +0.3 | 73.8 (67.9, 79.6) | 1.48 (1.38, 1.57) | +0.9 | 48.0 (40.8, 55.2) | 1.83 (1.67, 1.99) | +1.7 | 25.2 (18.4, 31.9) |
| 25 | 0.85 (0.82, 0.89) | +0.2 | 72.4 (66.7, 78.0) | 1.45 (1.36, 1.54) | +0.6 | 45.9 (39.3, 52.5) | 1.79 (1.64, 1.93) | +1.1 | 23.1 (17.2, 29.0) |
| 50 | 0.84 (0.80, 0.87) | 0.0 | 69.6 (64.1, 75.0) | 1.40 (1.31, 1.49) | 0.0 | 41.7 (36.0, 47.5) | 1.69 (1.57, 1.82) | 0.0 | 19.3 (14.5, 24.1) |
| 50.5 (median) | 0.84 (0.80, 0.87) | 0.0 (ref) | 69.5 (64.1, 75.0) | 1.40 (1.31, 1.48) | 0.0 (ref) | 41.6 (35.9, 47.4) | 1.69 (1.56, 1.82) | 0.0 (ref) | 19.2 (14.4, 24.1) |
| 100 | 0.80 (0.76, 0.84) | -0.4 | 63.3 (57.1, 69.6) | 1.29 (1.19, 1.39) | -1.3 | 33.3 (26.8, 39.7) | 1.50 (1.36, 1.65) | -2.3 | 12.6 (7.9, 17.3) |
| 200 | 0.71 (0.62, 0.80) | -1.5 | 48.5 (35.2, 61.8) | 1.03 (0.82, 1.25) | -4.4 | 17.5 (6.1, 28.9) | 1.12 (0.84, 1.41) | -6.8 | 3.8 (0.0, 8.5) |
| 378.6 (max) | 0.49 (0.26, 0.73) | -4.1 | 19.3 (0.0, 44.7) | 0.58 (0.19, 0.96) | -9.8 | 1.9 (0.0, 7.9) | 0.58 (0.18, 0.99) | -13.3 | 0.1 (0.0, 0.4) |
| **Danish centenarians (N = 128) — horizons 1, 2, 3 years** | | | | | | | | | |
| 22 (min) | 0.93 (0.90, 0.96) | +0.2 | 81.0 (74.5, 87.5) | 1.60 (1.50, 1.71) | +0.9 | 54.1 (45.1, 63.0) | 2.03 (1.85, 2.21) | +1.8 | 32.1 (23.4, 40.7) |
| 25 | 0.93 (0.90, 0.96) | +0.2 | 80.6 (74.1, 87.1) | 1.59 (1.49, 1.70) | +0.8 | 53.3 (44.4, 62.2) | 2.01 (1.83, 2.19) | +1.6 | 31.2 (22.8, 39.7) |
| 49.3 (median) | 0.91 (0.88, 0.95) | 0.0 (ref) | 77.1 (70.3, 83.9) | 1.53 (1.43, 1.63) | 0.0 (ref) | 46.8 (38.4, 55.1) | 1.88 (1.71, 2.04) | 0.0 (ref) | 24.6 (17.5, 31.6) |
| 50 | 0.91 (0.88, 0.95) | 0.0 | 77.0 (70.2, 83.8) | 1.53 (1.43, 1.63) | 0.0 | 46.6 (38.3, 54.9) | 1.87 (1.71, 2.04) | -0.1 | 24.4 (17.3, 31.4) |
| 100 | 0.88 (0.83, 0.92) | -0.4 | 68.1 (59.8, 76.4) | 1.37 (1.25, 1.48) | -1.9 | 32.5 (23.2, 41.8) | 1.58 (1.40, 1.76) | -3.6 | 12.5 (6.2, 18.8) |
| 200 | 0.76 (0.67, 0.85) | -1.8 | 43.4 (26.1, 60.8) | 0.99 (0.76, 1.22) | -6.5 | 8.8 (0.0, 19.2) | 1.03 (0.75, 1.31) | -10.2 | 1.1 (0.0, 3.6) |
| 460.9 (max) | 0.33 (0.13, 0.53) | -7.0 | 0.2 (0.0, 1.7) | 0.33 (0.12, 0.54) | -14.4 | 0.0 (0.0, 0.0) | 0.33 (0.12, 0.54) | -18.6 | 0.0 (0.0, 0.0) |
| **Danish nonagenarians (N = 176) — horizons 2, 4, 6 years** | | | | | | | | | |
| **Plasma NfL (pg/mL)** | **2 y: RMST (95% CI)** | **2 y: Δ (months)** | **2 y: S(τ), % (95% CI)** | **4 y: RMST (95% CI)** | **4 y: Δ (months)** | **4 y: S(τ), % (95% CI)** | **6 y: RMST (95% CI)** | **6 y: Δ (months)** | **6 y: S(τ), % (95% CI)** |
| 13.9 (min) | 1.79 (1.73, 1.86) | +0.2 | 77.0 (71.3, 82.7) | 3.04 (2.85, 3.23) | +1.0 | 48.0 (40.4, 55.7) | 3.76 (3.45, 4.08) | +1.9 | 25.6 (18.7, 32.4) |
| 25 | 1.78 (1.71, 1.85) | +0.1 | 75.8 (70.1, 81.5) | 2.99 (2.81, 3.18) | +0.5 | 46.0 (38.8, 53.3) | 3.68 (3.38, 3.97) | +0.8 | 23.6 (17.4, 29.9) |
| 34.1 (median) | 1.77 (1.70, 1.84) | 0.0 (ref) | 74.9 (69.1, 80.6) | 2.96 (2.77, 3.14) | 0.0 (ref) | 44.4 (37.4, 51.4) | 3.60 (3.32, 3.89) | 0.0 (ref) | 22.1 (16.2, 27.9) |
| 50 | 1.76 (1.68, 1.83) | -0.2 | 73.0 (67.2, 78.9) | 2.89 (2.70, 3.08) | -0.8 | 41.4 (34.6, 48.3) | 3.48 (3.20, 3.76) | -1.5 | 19.4 (13.9, 25.0) |
| 71.4 (P90) | 1.73 (1.65, 1.81) | -0.5 | 70.4 (64.1, 76.8) | 2.80 (2.59, 3.00) | -2.0 | 37.4 (30.2, 44.7) | 3.31 (3.01, 3.61) | -3.6 | 16.1 (10.5, 21.7) |
| 100 | 1.69 (1.60, 1.79) | -1.0 | 66.7 (58.9, 74.4) | 2.66 (2.41, 2.91) | -3.6 | 32.1 (23.5, 40.7) | 3.08 (2.70, 3.45) | -6.3 | 12.1 (5.9, 18.2) |
| 200 | 1.53 (1.32, 1.73) | -3.0 | 50.9 (33.8, 68.1) | 2.14 (1.61, 2.66) | -9.8 | 15.1 (1.3, 28.8) | 2.29 (1.59, 2.99) | -15.8 | 3.0 (0.0, 8.1) |
| 384.7 (max) | 1.07 (0.49, 1.65) | -8.4 | 17.7 (0.0, 48.8) | 1.19 (0.31, 2.06) | -21.2 | 0.8 (0.0, 4.6) | 1.19 (0.30, 2.09) | -28.9 | 0.0 (0.0, 0.1) |
| *Abbreviations*: CI, confidence interval; NfL, neurofilament light chain; RMST, restricted mean survival time; S(τ), covariate-adjusted probability of surviving to horizon τ. Predictions from flexible parametric survival models adjusted for age, sex and activities of daily living (Barthel Index in the Dutch, Katz Index in the Danish cohorts), at the cohort mean. Δ (months) is the RMST difference versus the cohort median NfL; positive values indicate longer survival. | | | | | | | | | |

| **Extended Data Table 9. Pre- and post-imputation descriptive statistics for MMSE and Barthel Index in Dutch centenarians** | | | | |
| --- | --- | --- | --- | --- |
| **Measure  (Theoretical Range)** | **Imputation Status** | **N (%)** | **Observed range (min, max)** | **Observed  Median (IQR)** |
| MMSE (0, 30) | Pre-imputation | 248/255 (97%) | 12, 30 | 26 (22, 28) |
|  | Post-imputation | 249/255 (98%) | 12, 30 | 26 (22, 28) |
| Barthel Index (0, 20) | Pre-imputation | 251/255 (98%) | 1, 20 | 15 (12, 18) |
|  | Post-imputation | 255/255 (100%) | 1, 20 | 15 (12, 18) |
| *Abbreviations*: BI, Barthel Index; IQR, interquartile range; MICE, multiple imputation with chained equations; MMSE, Mini-Mental State Examination. This imputation procedure was solely applied to the Dutch centenarian cohort. The MMSE measures global cognition by assessing orientation, memory, attention, language, and visuospatial skills. The BI measures independence in activities of daily living. Higher scores on both scales denote better performance. Item-level missing data caused by sensory or motor limitations were imputed using MICE (v3.13.0). Six participants with ≥6 missing MMSE items were excluded from the MMSE dataset. Assessments were administered on the day of blood sampling for 217 participants (85%) and within a median of 3.1 months (IQR 1.7–5.3) for the remaining 38 (15%). | | | | |

| **Extended Data Table 10. Interaction terms between plasma biomarkers and clinical function scores each estimated in a separate multivariable Cox model for all-cause mortality** | | |
| --- | --- | --- |
| **Predictor** | **HR (95% CI)** | ***P*-value** |
| **Dutch centenarians** |  |  |
| Aβ_42/40_ ratio × MMSE | 1.07 (0.93, 1.24) | 0.326 |
| Aβ_40_ × MMSE | 1.08 (0.93, 1.26) | 0.300 |
| Aβ_42_ × MMSE | 1.08 (0.94, 1.26) | 0.244 |
| pTau-181 × MMSE | 1.01 (0.86, 1.19) | 0.909 |
| pTau-181/Aβ_42_ ratio × MMSE | 0.96 (0.83, 1.11) | 0.581 |
| NfL × MMSE | 1.06 (0.95, 1.18) | 0.326 |
| GFAP × MMSE | 1.01 (0.90, 1.14) | 0.851 |
| Aβ_42/40_ ratio × BI | 1.14 (0.97, 1.33) | 0.102 |
| Aβ_40_ × BI | 0.91 (0.80, 1.05) | 0.196 |
| Aβ_42_ × BI | 0.96 (0.83, 1.12) | 0.607 |
| pTau-181 × BI | 0.99 (0.88, 1.10) | 0.803 |
| pTau-181/Aβ_42_ ratio × BI | 0.95 (0.85, 1.07) | 0.425 |
| NfL × BI | 1.02 (0.90, 1.16) | 0.771 |
| GFAP × BI | 0.99 (0.89, 1.10) | 0.824 |
| **Danish centenarians** |  |  |
| NfL × MMSE | 1.07 (0.87, 1.30) | 0.524 |
| NfL × Katz | 0.99 (0.78, 1.27) | 0.952 |
| **Danish nonagenarians** |  |  |
| NfL × MMSE | 0.88 (0.75, 1.03) | 0.101 |
| NfL × Katz | 0.99 (0.85, 1.15) | 0.881 |
| *Abbreviations*: Aβ, amyloid-β; BI, Barthel Index; CI, confidence interval; GFAP, glial fibrillary acidic protein; HR, hazard ratio; MMSE, Mini-Mental State Examination; NfL, neurofilament light chain; pTau-181, phosphorylated tau at threonine 181. Only the interaction HR and 95% CI are shown; main effects are omitted. No interaction reached *P*<0.05. Each model was restricted to participants with complete data on that interaction pair: Dutch centenarians N=249 for MMSE interactions (GFAP N=246) and N=255 for BI interactions (GFAP N=252), Danish centenarians N=128, and Danish nonagenarians N=176. | | |

Extended Data Figures

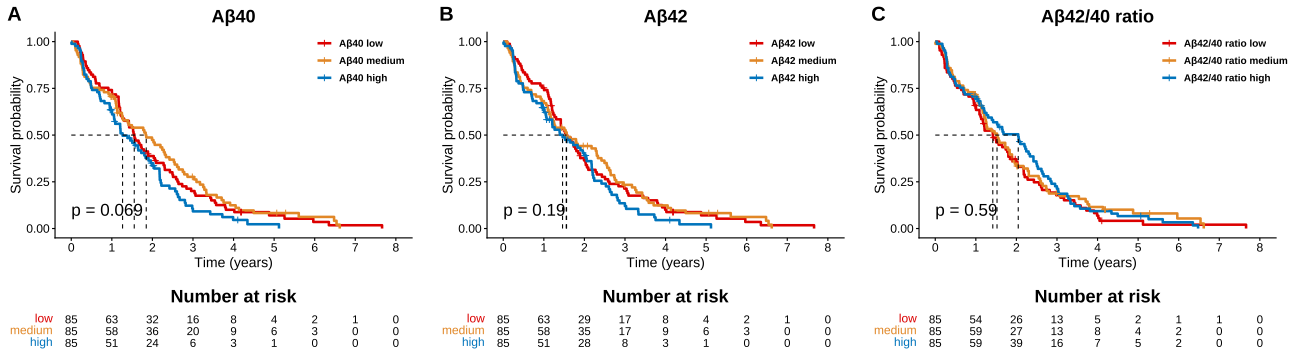

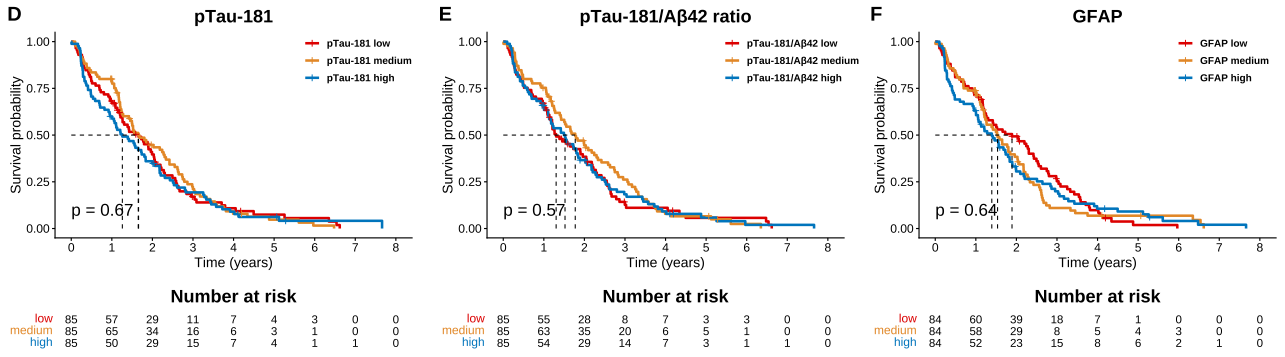

**Extended Data Figure 1. Kaplan-Meier survival curves for plasma biomarkers (except NfL) in Dutch centenarians.**

*Abbreviations*: Aβ, amyloid-β; BI, Barthel Index; GFAP, glial fibrillary acidic protein; MMSE, Mini-Mental State Examination; pTau-181, phosphorylated tau at threonine 181. Panels show cumulative survival probability (y-axis) from time of blood collection to death (x-axis, years) for participants classified into tertiles: low (red), medium (orange), and high (blue). (A–F) Plasma biomarker distributions for (A) Aβ_40_, (B) Aβ_42_, (C) Aβ_42/40_ ratio, (D) pTau-181, (E) pTau-181/Aβ_42_ ratio, and (F) GFAP. The *P*-value was derived from a log-rank test comparing survival distributions across the three tertile groups. Dashed lines indicate median survival times. Risk tables below each plot show the number of participants at risk over time. Sample sizes: Aβ biomarkers, pTau-181, pTau-181/Aβ_42_ ratio, N=255; GFAP, N=252.

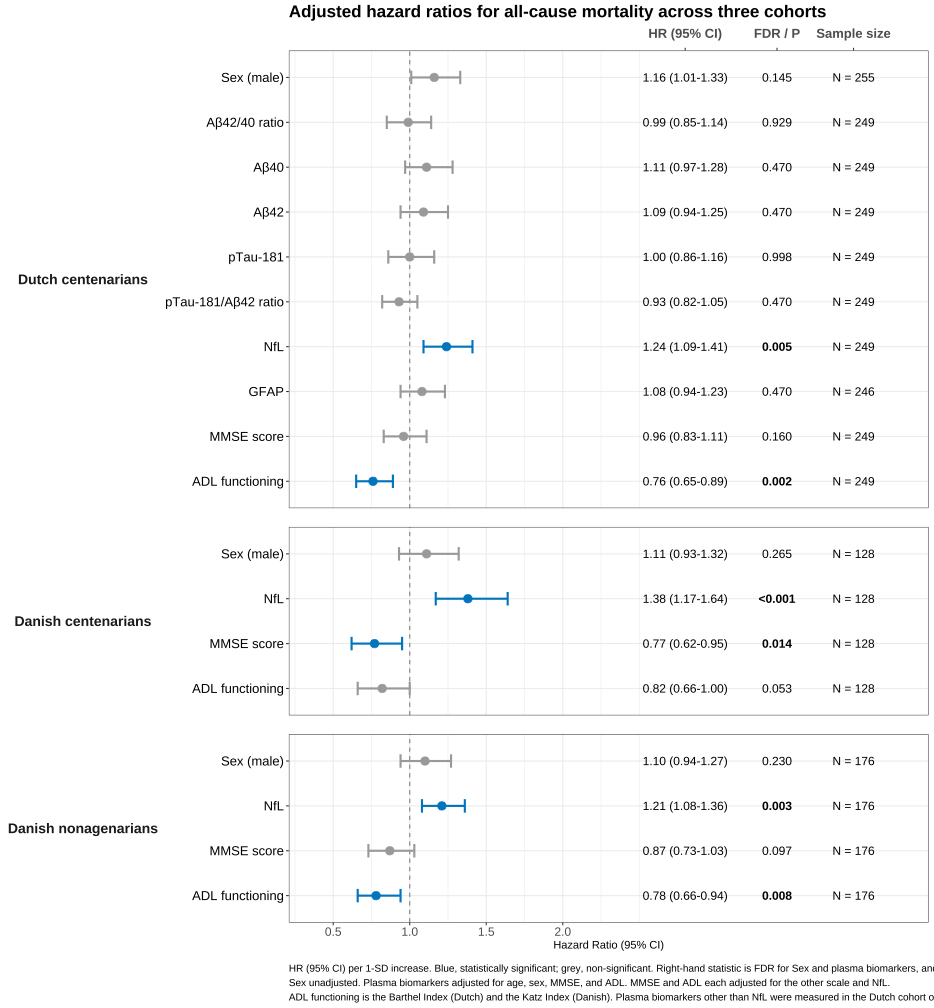

**Extended Data Figure 2. Forest plots of hazard ratios for all-cause mortality associated with demographics, plasma biomarkers and multi-item scales for cognitive and ADL physical functioning.**

*Abbreviations*: ADL, activities of daily living; Aβ, amyloid-β; BI, Barthel Index; CI, confidence interval; FDR, false discovery rate; GFAP, glial fibrillary acidic protein; HR, hazard ratio; Katz, Katz Index; MMSE, Mini-Mental State Examination; NfL, neurofilament light chain; pTau-181, phosphorylated tau-181; SD, standard deviation. HRs (95%CI) are per 1-SD increase from Cox models. Blue indicates statistical significance (*FDR*<0.05 for sex and plasma biomarkers; *P*<0.05 for the NfL-adjusted MMSE and ADL); grey indicates non-significant. Plasma biomarkers were adjusted for age, sex, MMSE, and ADL (BI in the Dutch cohort, Katz in the Danish cohorts); MMSE was adjusted for age, sex, ADL, and NfL; ADL was adjusted for age, sex, MMSE, and NfL; Sex (male, with female as the reference) is shown unadjusted but is included in the FDR correction.

**Dutch centenarians (N=255)**

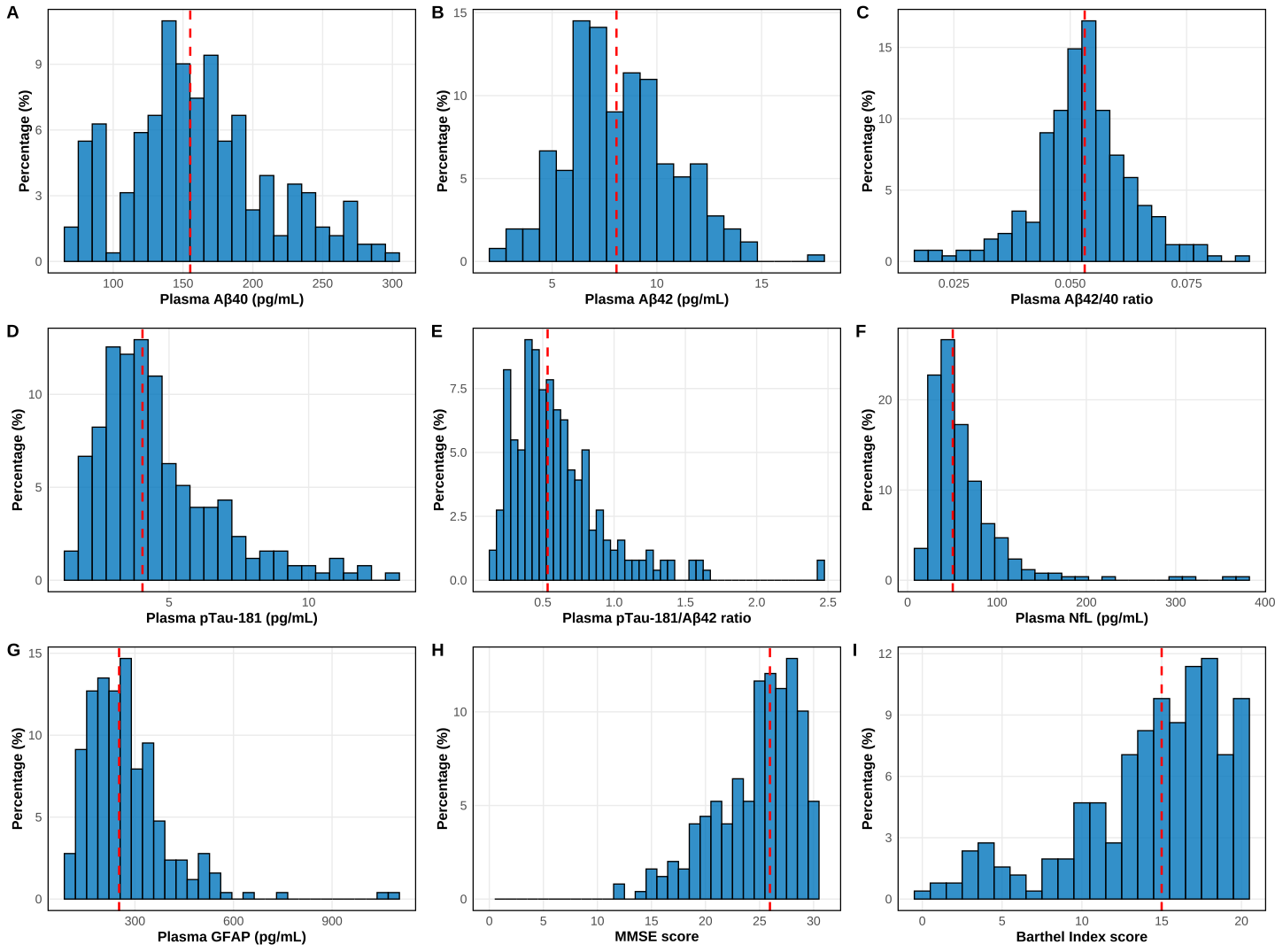

**Danish centenarians (N=128)**

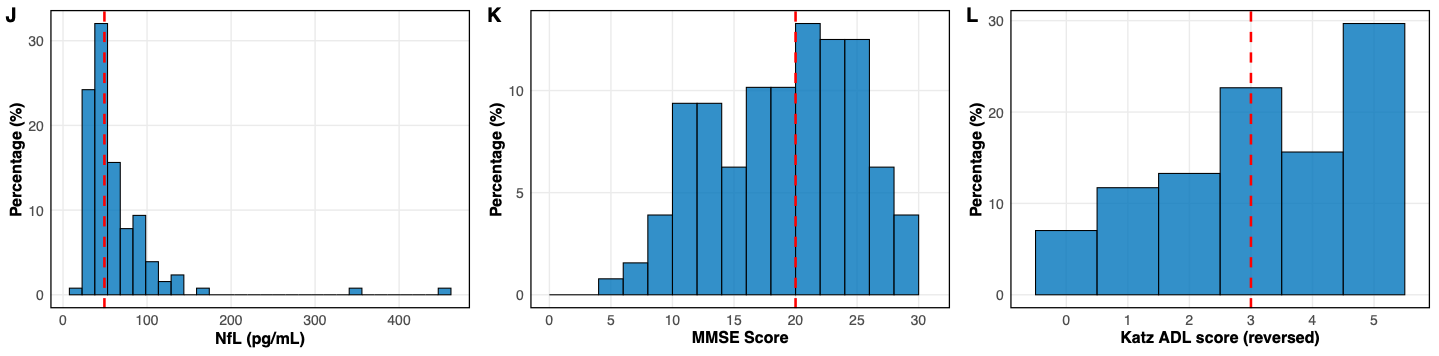

**Danish nonagenarians (N=176)**

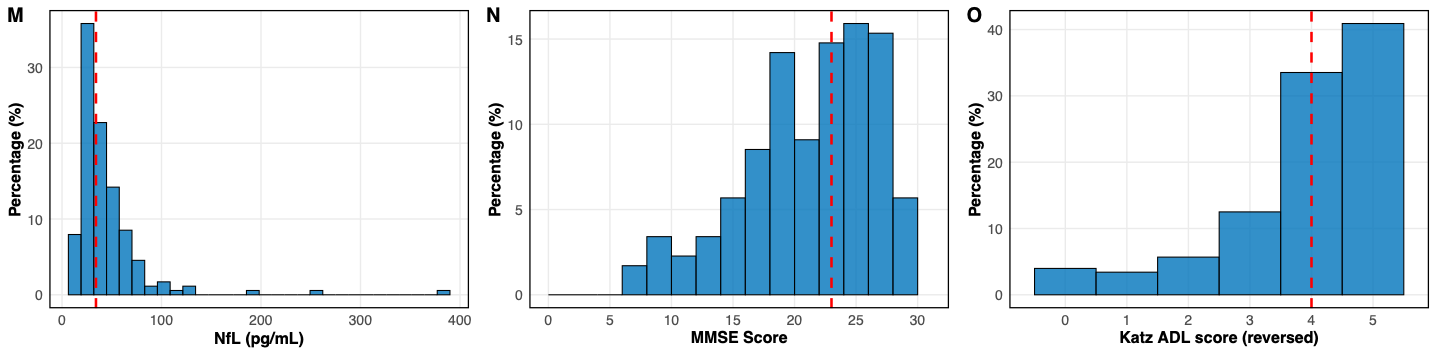

**Extended Data Figure 3.** **Distributions of plasma biomarkers, MMSE scores and ADL scores.** *Abbreviations*: Aβ, amyloid-β; BI, Barthel Index; GFAP, glial fibrillary acidic protein; MMSE, Mini-Mental State Examination; NfL, neurofilament light chain; pTau-181, phosphorylated tau at threonine 181. (A–I) Histograms show plasma concentrations of (A) amyloid-β40 (Aβ_40_), (B) amyloid-β42 (Aβ_42_), (C) Aβ_42/40_ ratio, (D) phosphorylated tau at threonine 181 (pTau-181), (E) pTau-181/Aβ_42_ ratio, (F) neurofilament light chain (NfL), and (G) glial fibrillary acidic protein (GFAP), and clinical score distributions for (H) Mini-Mental State Examination (MMSE) and (I) Barthel Index in Dutch centenarians. (J–L) Histograms show distributions of (J) NfL, (K) MMSE, and (L) Katz ADL score in Danish centenarians. (M–O) Histograms show distributions of (M) NfL, (N) MMSE, and (O) Katz ADL score in Danish nonagenarians. Red dashed lines indicate the median value for each distribution. Sample sizes: Dutch centenarians (Aβ_40_, Aβ_42_, Aβ_42/40_ ratio, pTau-181, pTau-181/Aβ_42_ ratio, and NfL, N=255; GFAP, N=252; MMSE, N=249; Barthel Index, N=255); Danish centenarians (N=128); and Danish nonagenarians (N=176).

14	powerSurvEpi: Power and Sample Size Calculation for Survival Analysis of Epidemiological Studies v. 0.1.5 (Comprehensive R Archive Network (CRAN), 2025).

15	survSNP: Power Calculations for SNP Studies with Censored Outcomes v. 0.26 (Comprehensive R Archive Network (CRAN), 2023).
